# Breaking Barriers: A cross-sectional study on Menstrual Restrictions and Perceived Stress among adolescent girls in Kailali, Nepal

**DOI:** 10.1101/2023.03.31.23287836

**Authors:** Alisha Dahal, Krishna Prasad Sapkota, Deepa Kumari Bhatta, Ankit Acharya

## Abstract

Menstrual restriction has persisted in Nepalese society for centuries, driven by ignorance and myth. This practice is imposed on women through various Hindu mythologies and leads to significant limitations in their daily activities. The most severe form of menstrual restriction, known as Chaupadi, has resulted in numerous deaths of women and young children due to suffocation, snakebite, rape, and other serious forms of harm. Despite its criminalization by law, there has been no visible impact on the practice of menstrual restriction. This discriminatory practice not only causes hormonal imbalance and physical pain but also has significant mental health implications for adolescent girls, which have yet to be fully explored. Therefore, this study aims to investigate the prevalence and perceived stress associated with menstrual restriction among adolescent girls attending lower secondary school in Far-Western Region, Kailali District, Nepal

The study utilized a descriptive, cross-sectional design and recruited 370 respondents using a proportionate random sampling technique. The study was conducted at the secondary school of Godawari Municipality in Kailali, Nepal. Prior to the study, a structured questionnaire and a Likert scale were pretested among 10% of the population in Kathmandu. Data Analysis was done using Univariate Analysis, Bivariate Analysis (Chi-square) and Multi-variate Analysis (Logistic Regression).

The findings revealed that perceived stress was comparatively higher among upper-caste groups and Dalits who followed menstrual restrictions religiously, as compared to disadvantaged Janajati from the hills. Moreover, the level of perceived stress was moderately high among households with larger family sizes. Significant associations were observed between menstrual restriction and perceived stress for each type of menstrual restriction studied, with more than 80% of the different measured levels of restrictions showing a significant association. Menstrual restriction-related practices, such as being restricted from entering inside the house, being sent to Chau-Goth, being restricted from touching male members, being restricted from touching livestock or animals, being restricted from eating together with the family, being restricted from consuming dairy products, being restricted from participating in cultural rituals, being restricted from sleeping in any bed, and being restricted from using heavy blankets and mattresses, were associated with perceived stress. The relationship between menstrual restrictions and perceived stress level was examined. Results showed that respondents with restrictions to enter or reside inside the house had 3.78 times higher odds of perceived stress (OR=3.78; CI=1.96-7.33), while those sent to Chhau Goth had 2.98 times higher odds (OR=2.98; CI=1.94-4.57). Respondents with restrictions on touching food, cooking food, plants with holy belief, livestock or animals, dairy products, eating together with family, participating in cultural rituals, having Prasad, sleeping in any bed, using packed pads, common toilet, or common taps during menstruation also had higher odds of perceived stress. Given the findings, further research is necessary to measure the level of perceived stress among adolescent girls in the population. This research has significant implications for the physical, social, and psychological well-being of adolescent girls and the community at large.

## Introduction

"Menstruation marks a crucial period in an adolescent’s life, characterized by profound physical, mental, and psychological changes."(1) "For adolescent girls, the onset of their first menstruation cycle signifies a pivotal transition from childhood to womanhood." (2) However in Nepal, the practice of "Chaupadi," in which women are isolated to small huts or cow sheds during menstruation, has long been a cultural and traditional taboo. Despite efforts to eradicate the practice, including a Supreme Court ban in 2004 and a 2010 recognition by the National Plan of Action against Gender-Based Violence that it constitutes a form of violence against women, Chaupadi persists in some rural areas of Nepal. The Nepalese parliament passed a bill in 2018 aimed at punishing anyone who practices Chaupadi, but the practice still endures in the far-western and mid-western regions, particularly among families who follow Hindu religion.

In Nepal, menstruation is widely regarded as a social taboo due to poor knowledge and information about reproductive health, particularly in rural areas. Adolescent girls who experience menstruation are subjected to taboo practices that are deeply rooted in Nepali culture and traditions. During this time, women are prohibited from worshipping, entering the kitchen, cooking food for their family, and even staying at home. Additionally, menstruating women are considered impure and are not allowed to touch anyone. Although cultural norms have shifted in some western settings, these taboos still persist in many rural areas of Nepal. (2–4)Young adolescent girls are at high risk of experiencing various degrees of stress during puberty. When cultural and traditional taboos accompany these changes, it can lead to poor mental and physical health. Unfortunately, stigma and discrimination are still prevalent in Nepal. In the hilly areas of provinces 6 and 7, women are forced to spend their menstrual period in a small hut or cow shed, known as "Chaupadi". Women and girls who follow this traditional taboo face harassment, sexual abuse, rape, fear of wild animals, isolation from family members, and are regarded as untouchable. This leads to mental stress, frustration, trauma, and depression, and raises serious questions about women’s safety. (5)

Stress can be defined as a normal reaction which affects our mental, emotional or physical state and ultimately results in negative health problems such as depression, anxiety etc. It is known to be one of the crucial factors that play a major role in developing depression, cardio-vascular disease or immune related disorders as well. (6) While menstrual restrictions, including Chaupadi, have long been a public health concern in Nepal, little research has been done to study their impact on the mental health of adolescent girls. This study aims to fill that gap by exploring the association between menstrual restriction and perceived stress among young girls in Nepal. This study aims to investigate the impact of menstrual restriction on the mental health of young adolescent girls in Nepal, particularly focusing on the practice of Chaupadi. By collecting relevant data, this research can provide evidence to design interventions that eliminate social taboos and superstitious beliefs related to menstrual restriction. The study’s results can provide evidence to sensitize and motivate government policies and programs to address the mental health hazards caused by menstrual restrictions.

## Objective

### General objective

To access the prevalence and perceived stress caused due to menstrual restriction among Adolescent girls of Lower Secondary School in Kailali District, Nepal.

### Specific objective

1. To measure the prevalence of different forms of menstrual restriction.
2. To measure the Perceives Stress level of Adolescent girls due to menstrual restriction.
3. To measure the association between menstrual restriction and perceived stress.

### Research Hypothesis

**Ho:** There is no any association between perceived stress level and menstrual restriction

**H1:** There is significant association between perceived stress level and menstrual restriction

## Methods

### Study Design

The study applied a descriptive analytical cross-sectional study design using a self-administerd questionnaire.

### Research setting

Kailali District lies in the far western region/Province 7 of Nepal. A region where restriction practice in regards to Menstruation is found in each and every family member. (7,8) Even though the Nepal government played an important role to ban “Chaupadi pratha” in 2005 by taking certain action, it is still in practice. (9) Thus there is a high risk of women and girls who have to face these social taboos.

The most extreme of these practices is the ‘Chhaupadi Pratha’ (Chhaupadi), which is prevalent throughout the far western region and some parts of the mid-western region of Nepal (10) people living in Kailali district in Province Number Seven as opposed to Bardiya district in Province Number Five had significantly higher reproductive health problems such as burning during micturition, abnormal discharge, genital itching, or painful and foul smelling. (11)

The study was conducted in Government Schools of State-7, Godawari Municipality 10, 11 of Kailali district, Nepal.The sample students were enrolled from 3 different schools i.e. Shree Gwasi Secondary school, Shree Saraswati Secondary School, Shree Janaki Secondary School. In Shree Gwasi Secondary School.

### Participant

In total 370 students participated in the study.This research is mainly focused to assess perceived stress related to menstrual restrictions among the adolescent girls of age 14 to 17 years in Kailali District of Nepal. The age group was chosen based on the grade they would be studying and to access the perceived stress level of girls form late adolescence as age of menarche may differ in lower grades.

The sample students were enrolled from the adolescent girls who were studying in 8, 9 and 10 grade of government schools of Godawari municipality, Kailali district in Sudurpaschim Province, Nepal.

Among 36 schools present in the Municipality, there were 6 secondary level schools among which 3 schools were located in the Godavari Municipality which were selected randomly. The 3 schools were taken in the sample as Shree Gwasi Secondary School (Ward no 3), Shree Saraswati Secondary School (Ward 11) and Shree Janaki Secondary School (Ward 12). After the approval from the Nepal Health Research Council on February 17^th^ , 2019 ; data collection was started from the 4^th^ week of February until 1^st^ week of March. The study period was from 4^th^ week of February to 4^th^ week of June, 2019.

Questionnaires were distributed by the researcher with verbal instruction to each participant adolescent girls who were present during the data collection. The name list of all the adolescent girls were prepared coordinating with the school administration and random sampling was performed to get the ideal sample from each class. The checklist was prepared to keep track record of questionnaire distributed and responses collected. Each participant were given a reference number in the form they had filled, this way the researcher did not know about the identity of the participant and it was kept anonymous.

### Criteria for sample selection

1. **Inclusion:**

- Female students of Class 8, 9, 10 in Governmental School.
2. **Exclusion:**

- Absentees
- Other students than of the taken Municipality

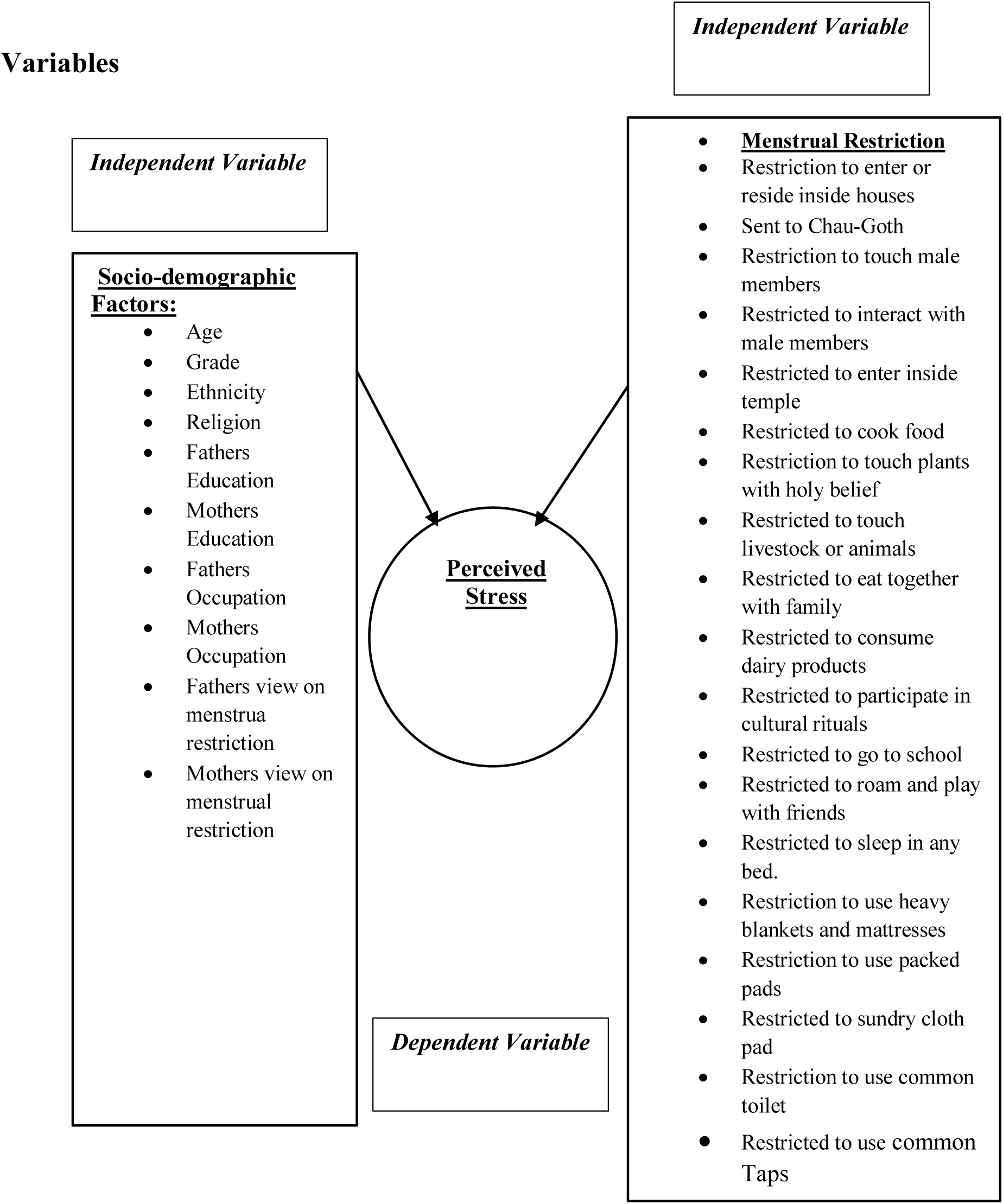

### Data Source

#### Validity and reliability of tool

The validity of the instruments was ascertained by consulting with the adviser, expertise, literature review and discussion with supervisor and by pretesting results of the instrument on 10 % of total sample size in Kailali. The items relevant to the Nepalese context was adopted based on A Global Measure of Perceived Stress (PSS) by Sheldon Cohen, Tom Kamarck, and Robin Mermelstein which are valid scales and have a high reliability (Coefficient batch alpha 0.84, 0.85, 0.86 respectively,(12) which was also previously use in the study done in 2003, in the title “ Psychological Factors in Nepali Former Commercial Sex workers with HIV” in Nepal. (13) A total of 10 item measure (Table 1)of the degree of which life situations are appraised in a five point Likert Scale. Respondents were asked to indicate about their experience in stress related feeling and thoughts on a 5-point scale as 0=never, 1=almost never, 2=sometimes, 3=fairly often, 4=very often. (12,13)

Validity of the instruments was ascertained through Chronbatch alpha for both Perceived Stress Scale and Menstrual Restriction to measure the internal consistency. The Chronbatch alpha for both was found to be 0.82 for PSS-10 Scale and 0.85 for menstrual restriction. Expert consultation, literature review, and the questionnaire was piloted among 10% adolescents with similar characteristics as the study population in a similar Far-Western setting. The purpose of the pre-test was to ascertain the clarity, sensitivity, and practicability of the questionnaire and to identify ambiguous and poorly constructed items as well as other problems that may be encountered during data collection.

A structured self-administered questionnaire was developed based on intensive literature review. The whole questionnaire was translated in Nepali and back translation was done into English before pretesting. Pretesting was carried out among 10% of the total population**.(Annex 1)**

### Tools and technique

#### Structured questionnaire

Self-administered questionnaire in nepali language including 3 parts and a participant information sheet with an informed conscent form was used. First part of the questionnaire included questions related sociodemographic information. The second part of the tool consisted information about various types of menstrual restriction practiced in Nepal through thorough literature review, expert advise and supervision from the supervisor. The third part of the questionnaire was adopted from the perceived stress scale (PSS).

#### Perceived Stress Scale

Perceived Stress Scale is known as a classic stress assessment instrument which is based on A Global Measure of Perceived Stress (PSS) by Sheldon Cohen, Tom Lamarck, and Robin Mermelstein, which are valid scales and have a high reliability (Coefficient batch alpha 0.84, 0.85, 0.86 respectively). The 10 items in PSS were relevant in Nepalese context which measure the degree of an individual’s situation is evaluated as stressful. (12) While using this tool 5 point Likert scale were applied where 0=never, 1=almost never, 2=sometimes, 3=fairly often, 4=very often respectively. The total score of PSS was then calculated by adding the sum of all 10 items that were used. (13)

In regards to validity, Cronbach alpha was used for both perceived stress scale and menstrual restriction to measure the internal consistency. The value of Cronbach alpha was found to be 0.82 and 0.85 for PSS-10 scale and menstrual restriction respectively which was calculated using the SPSS Vs. 24. Pretesting was conducted in 10 % of the total sample size in Kathmandu valley to check the reliability of the study.

#### Bias

Recall bias is one of the potential bias in the study, as girls who will not be going through menstruation at the moment must go back to think. Since its self administered there is some bias too. We tried to manage this bias by trying to make them recall the event during the orientation of the questionnaire and constantly probing that these questions were asking about the time of their last or current menstruation.

### Study Size

#### Sample Size and Sampling

A sample size of 308 was estimated using the following simple formula to calculate the adequate sample size in prevalence study, *n*=*Z*2*P*(1−*P*)*d*2 (14) , Where n is the sample size, Z is the statistic corresponding to level of confidence which is 95%, P is expected prevalence of 72.3 prevalence of menstrual restriction prevalence from a previous study (15)and d is precision (corresponding to effect size) of 5%.

Sample size was calculated by using the following formula

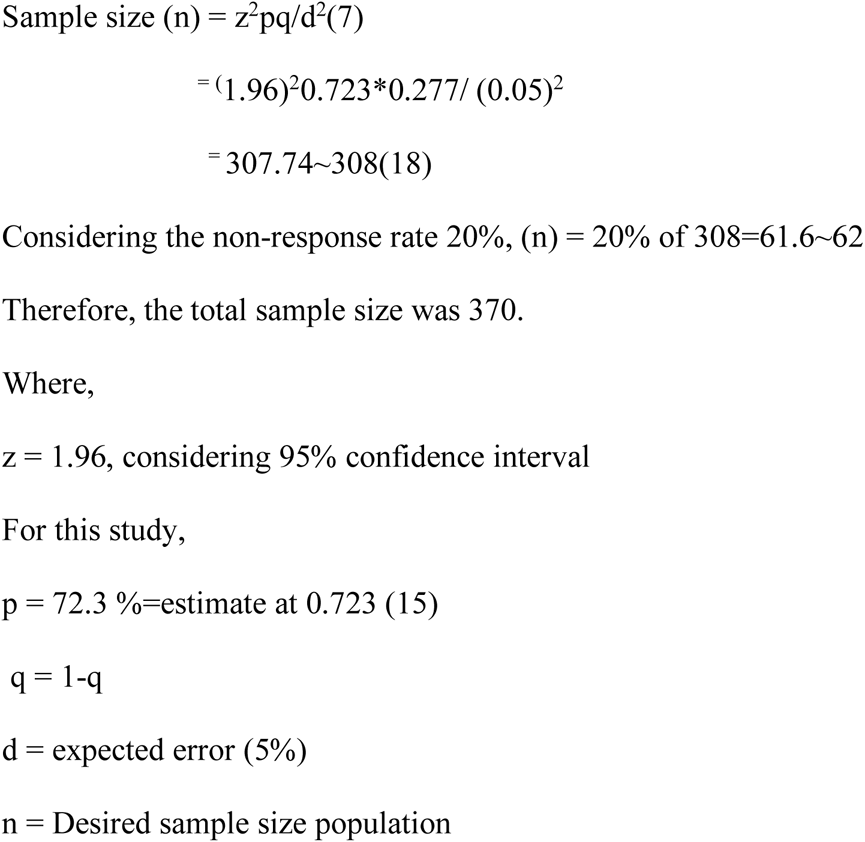

Upon approval from the Godawari Municipality we got the list of the schools and approval form the school heads of the randomly allocated 3 governent schools f secondary level. Provided the list then we adopted Proportionate random sampling technique for this research as shown belowin **Figure 1**. To draw the samples, firstly three schools were randomly selected, and then total no. of students was identified from the selected schools. No of samples from each class (8, 9, and 10) from each school were calculated proportionately and students were selected randomly from each class.

**Figure 1.**
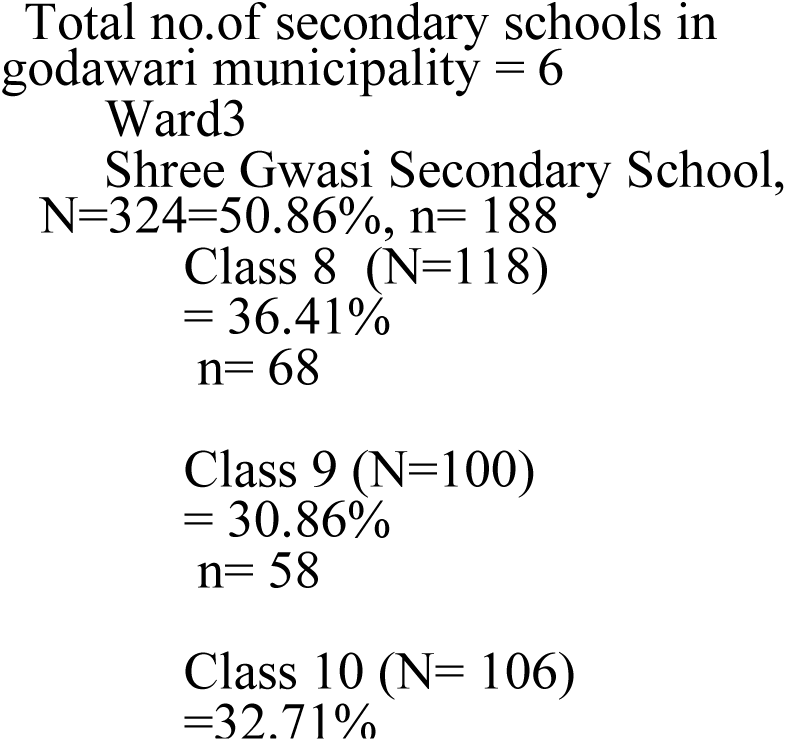
Proportionate random Sampling Steps

### Quantitaive Variables

The received questionnaires were rechecked for any missing data, followed up and filled then coded before transfer of the data. Immediately following the data collection, prior to data entry, data was checked for completeness. The researcher managed and transferred the filled questionnaire own self to the destination. Researcher screened all the data for accuracy, assurance and completeness. This promptness helped to address any omissions, errors or inaccuracies also helping to correct the data. Then finally, the responses were transferred into computer software Epi-Data to create a data set. Data were stored in specific folder of laptop and backup was stored in external hard disk for prevention from loss.

EpiData v3.1 was used for data entry. 10% raw data was entered after developing format for validating the data entry format. At the end of the data entry 10% of filled questionnaires was selected randomly and crosschecked with the entered data.The original data set was kept safely in a separate folder to prevent from manipulation and loss. Recoding and analysis was conducted in a copy of the data set in a separate folder.

First, data was entered in EpiData v3.1 and then exported to SPSS for analysis. The data wasthen analyzed by using the software SPSS v24.0 (Statistical Package for Social Sciences), referencing was done using Mendeley Software for Report Writing purpose. After the entry of data the questionnaires were packed properly and has been kept in the dark room in the primary investigators store house.

### Statistical Methods

Since, the missing data were handled during the data collection process there were not any missing data in this study. Analyzed data was presented in tables and graphs in the descriptive section presenting the count, percentage and cross-tabulation. In the second part, Bivariate analysis was conducted between Socio-demographic, Menstrual Restriction and Perceived Stress. For the interpretation of the data P value at 95% confidence interval was used with the help of Chi-square test. All data were interpreted based on p-value <0.05 being significantly associated and p-value >0.05 being not significant. Further, The associated variables were then used to see the effect size using the Logistic regression method and were calculated in Adjusted ODDs ratio using Multi Variate Analysis. All the referencing for the manuscript was done using Mendeley Software.

### Ethics statement

Research permits for the study were received by different concerned authorities. Ethical clearance and approval was obtained from the Ethics and review committee of Nepal Health Research Council (NHRC), Ref : 2274.

The survey was conducted in 3 different government schools of Godavari Municipality of Kailali district. Permission was obtained from respective schools and municipalities as well in order to collect data in school premises. Likewise, written consent was obtained from each of the school’s headmaster or headmistress for their school’s participation in the study.

Furthermore, the students signed written consent forms as well as assent forms in Nepali (Language spoken by most of the students in the schools of Kailai districts is Nepali) prior to data collection. The form explains the objective or purpose of this study along with its risks and significance of the study. The form clearly states participants’ rights to participate or withdraw. Students’ parents and guardians were informed about the research and its purpose through school authority. Parents were provided with a written conscent prior to data collection activity.

The studys objective and the questionnaire were explained to all the participants and they were assured of their anonymity and the confidentiality of their responses.

## Results

### Univariate Analysis: Socio-demographic variables of the participants

**Table 1.**
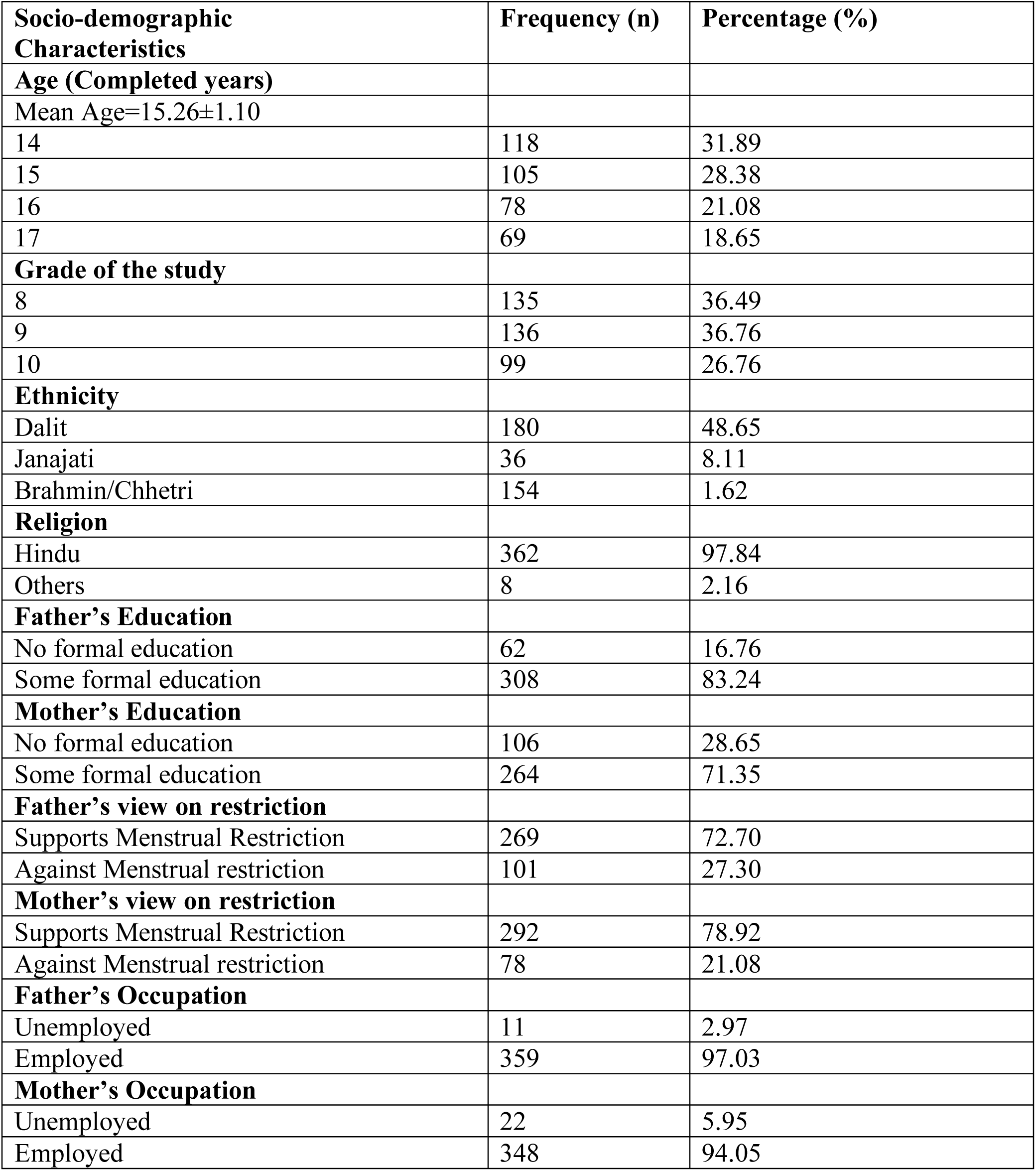
Univariate Analysis of socio-demographic variable, n=370

Out of the 370 participants, the mean age was 15.26±1.10 years. The distribution of participants across grades was almost equal with 36.49% in grade 8, 36.76% in grade 9, and 26.76% in grade 10. Among the participants, 48.65% were Dalit, 8.11% were Janajati, and 1.62% were Brahmin/Chhetri. Nearly all participants (97.84%) identified as Hindu. The majority of the fathers (83.24%) had some level of formal education, while 28.65% of the mothers had no formal education. The majority of both fathers (72.70%) and mothers (78.92%) supported menstrual restriction. Most of the mothers (94.05%) and almost all of the fathers (97.03%) were employed.

### Bi-variate Analysis: Sociodemographic Variable and Perceived Stress

**Table 2.** The relationships between demographic variables of the participants and the perceived stress, n=370

| **Socio-demographic Characteristics** | **Total** | **High PS\*** | **Low PS\*** | **$\chi^2$ Value** | **P-Value** |
| --- | --- | --- | --- | --- | --- |
| **Age (Completed years)** |  |  |  | 8.82 | **0.03** |
| Mean Age=15.26±1.10 |  |  |  |  |  |
| 14 | 118 | 54(45.76) | 64(54.24) |  |  |
| 15 | 105 | 44(41.90) | 61(58.10) |  |  |
| 16 | 78 | 49(62.82) | 29(37.18) |  |  |
| 17 | 69 | 36(52.17) | 33(47.83) |  |  |
| **Grade of the study** |  |  |  | 6.42 | **0.04** |
| 8 | 135 | 78(57.78) | 57(42.22) |  |  |
| 9 | 136 | 58(42.65) | 78(57.35) |  |  |
| 10 | 99 | 47(47.47) | 52(52.53) |  |  |
| **Ethnicity** |  |  |  | 20.99 | **0.001** |
| Dalit | 180 | 100(55.56) | 80(44.44) |  |  |
| Janajati | 36 | 5(13.89) | 31(86.11) |  |  |
| Brahmin/Chhetri | 154 | 78 (50.65) | 76(49.35) |  |  |
| **Religion** |  |  |  | 1.09 | 0.29 |
| Hindu | 362 | 181(50.00) | 181(50.00) |  |  |
| Others | 8 | 2(25.00) | 6(75.00) |  |  |
| **Father's Education** |  |  |  | 1.81 | 0.18 |
| No formal education | 62 | 36(58.06) | 26(41.94) |  |  |
| Some formal education | 308 | 147(47.73) | 161(52.27) |  |  |
| **Mother's Education** |  |  |  | 2.65 | 0.10 |
| No formal education | 106 | 60(56.60) | 46(43.40) |  |  |
| Some formal education | 264 | 123(46.59) | 141(53.41) |  |  |
| **Father's view on restriction** |  |  |  |  |  |
| Supports Menstrual Restriction | 269 | 133(49.44) | 136(50.56) | 0.00 | 0.99 |
| Against restriction | 101 | 50(49.50) | 51(50.50) |  |  |
| **Mother's view on restriction** |  |  |  |  |  |
| Supports Menstrual Restriction | 292 | 145(49.66) | 147(50.34) | 0.02 | 0.98 |
| Against Menstrual restriction | 78 | 38(48.72) | 40(51.28) |  |  |
| **Father's Occupation** |  |  |  |  |  |
| Unemployed | 11 | 9(81.82) | 2(18.18) | 3.51 | 0.06 |
| Employed | 359 | 174(48.47) | 185(51.53) |  |  |
| **Mother's Occupation** |  |  |  |  |  |
| Unemployed | 22 | 11(50.00) | 11(50.00) | 0.003 | 0.96 |
| Employed | 348 | 172(49.43) | 176(50.57) |  |  |

Data provided as n (%)
\*Percentages refer to rows (values in rows add up to 100%)
Level of significance=0.05; p-value came from $\chi^2$ test

Table 2 The chi square test results showed a significant correlation between age (2=8.82; p=0.03), study grade (2=6.42; p=0.04), and ethnicity (2=20.99; p=0.001) and perceived stress level. However, no significant association was found between perceived stress level and religion (χ2=1.09; p=0.29), father’s education (χ2=1.81; p=0.18), mother’s education (χ2=2.65; p=0.10), father’s view on restriction (χ2=0.00; p=0.99), mother’s view on restriction (χ2=0.02; p=0.98), father’s occupation (χ2=3.51; p=0.06), and mother’s occupation (χ2=0.003; p=0.96).

### Bi-variate Analysis: Types of Menstrual Restriction and Perceived Stress

**Table 3.** Association of menstrual restrictions with perceived stress level among school going adolescent girls, n=370

| Types of Restrictions | High PS* | Low PS* | $\chi^2$<br>Value | P-Value |
| --- | --- | --- | --- | --- |
| **Restricted to enter the inside house**<br>Yes (315)<br>No (55) | 170 (53.97%)<br>13 (23.64%) | 145 (46.03%)<br>42 (76.36%) | 16.04 | **0.001** |
| **Sent to Chau Goth**<br>Yes (210)<br>No (160) | 128 (60.95%)<br>55 (34.38%) | 82 (39.05%)<br>105 (65.63%) | 24.60 | **0.001** |
| **Restricted to interact with male**<br>Yes (71)<br>No (299) | 42 (59.15%)<br>141 (47.16%) | 29 (40.85%)<br>158 (52.84%) | 3.30 | 0.09 |
| **Restricted to touch male members**<br>Yes (94)<br>No (276) | 46 (48.94%)<br>137 (49.64%) | 48 (51.06%)<br>139 (50.36%) | 0.014 | 0.91 |
| **Restricted to cook food**<br>Yes (318)<br>No (52) | 169 (53.14%)<br>14 (26.92%) | 149 (46.86%)<br>38 (73.08%) | 11.26 | **0.001** |
| **Restricted to touch food to be eaten by others**<br>Yes (322)<br>No (48) | 169 (52.48%)<br>14 (29.17) | 153 (47.52%)<br>34 (70.83%) | 8.17 | **0.004** |
| **Restricted to eat together with a family**<br>Yes (272)<br>No (98) | 150 (55.15%)<br>33 (33.67%) | 122 (44.85%)<br>65 (66.33%) | 12.44 | **0.001** |
| **Restricted to touch livestock or animals**<br>Yes (216)<br>No (154) | 130 (60.19%)<br>53 (34.42%) | 86 (39.81%)<br>101 (65.58%) | 22.86 | **0.001** |
| **Restricted to consume dairy Products**<br>Yes (271)<br>No (99) | 152 (56.09%)<br>31 (31.31%) | 119 (43.91%)<br>68 (68.69%) | 16.83 | **0.001** |
| **Restricted to consume meat products**<br>Yes (53)<br>No (317) | 26 (49.06%)<br>157 (49.53%) | 27 (50.94%)<br>160 (50.47%) | 0.004 | 0.95 |
| **Restricted to consume vegetables**<br>Yes (52)<br>No (318) | 26 (50.00%)<br>157 (49.37%) | 26 (50.00%)<br>161 (50.63%) | 0.007 | 0.93 |
| **Restricted to consume fruits**<br>Yes (60)<br>No (310) | 28 (46.67%)<br>155 (50.00%) | 32 (53.33%)<br>155 (50.00%) | 0.11 | 0.74 |
| **Restricted to enter inside temple**<br>Yes (336)<br>No (334) | 171 (50.89%)<br>12 (35.29%) | 165 (49.11%)<br>22 (64.71%) | 2.41 | 0.12 |
| **Restricted to touch holy plants (Tulasi, Peepal etc.)**<br>Yes (312)<br>No (58) | 162 (51.92%)<br>21 (36.21%) | 150 (48.08%)<br>37 (63.79%) | 4.22 | **0.04** |
| **Restricted to participate in cultural rituals**<br>Yes (177)<br>No (193) | 99 (55.93%)<br>84 (43.52%) | 78 (44.07%)<br>109 (56.48%) | 5.20 | **0.02** |
| **Restricted to have *Prasad***<br>Yes (22)<br>No (348) | 17 (77.27%)<br>166 (47.70%) | 5 (22.73%)<br>182 (52.30%) | 6.10 | **0.01** |
| **Restricted to go to school**<br>Yes (38)<br>No (332) | 23 (60.53%)<br>160 (48.19%) | 15 (39.47%)<br>172 (51.81%) | 1.61 | 0.20 |
| **Restricted to roam around and play with friends**<br>Yes (61)<br>No (309) | 34 (55.74%)<br>149 (48.22%) | 27 (44.26%)<br>160 (51.78%) | 0.87 | 0.35 |
| **Restricted to sleep in usual bed**<br>Yes (315)<br>No (55) | 170 (53.97%)<br>13 (23.64%) | 145 (46.03%)<br>42 (76.36%) | 16.04 | **0.001** |
| **Restricted to sleep in any bed**<br>Yes (190)<br>No (180) | 114 (60.00%)<br>69 (38.33%) | 76 (40.00%)<br>111 (61.67%) | 16.50 | **0.001** |
| **Restricted to use heavy blankets and mattresses**<br>Yes (122)<br>No (248) | 68 (55.74%)<br>115 (46.37%) | 54 (44.26%)<br>133 (53.63%) | 2.51 | 0.11 |
| **Restricted to use packed pads**<br>Yes (72)<br>No (298) | 45 (62.50%)<br>138 (46.31%) | 27 (37.50%)<br>160 (53.69%) | 5.45 | **0.02** |
| **Restricted to use sundry cloth pads**<br>Yes (74)<br>No (296) | 42 (56.76%)<br>141 (47.64%) | 32 (43.24%)<br>155 (52.36%) | 1.62 | 0.20 |
| **Restricted to use the common toilet** |  |  |  |  |
| Yes (153) | 101 (66.01%) | 52 (33.99%) | 27.48 | **0.001** |
| No (217) | 82 (37.79%) | 135 (62.21%) |  |  |
| **Restricted to use common tap** |  |  |  |  |
| Yes (295) | 162 (54.92%) | 133 (45.08%) | 16.27 | **0.001** |
| No (75) | 21 (28.00%) | 54 (72.00%) |  |  |

Data provided as n (%) , \*Percentages refer to rows (values in rows add up to 100%), Level of significance=0.05; p-value came from $\chi^2$ test

Table 3 revealed a significant association between perceived stress during menstrual periods and several restrictions faced by adolescent girls. These restrictions included being prohibited from entering or residing inside the house (χ2 =16.04; p=0.001), being sent to Chhau Goth (χ2=24.60; p=0.001), cooking food (χ2=11.26; p=0.001), touching food to be eaten by others (χ2=8.17), eating together with family (χ2=12.44; p=0.001), touching live stocks (χ2=22.86; p=0.001), and consuming dairy products (χ2=16.83; p=0.001). Furthermore, the frequencies and p-values indicated that the perceived stress level was significantly associated with restrictions such as touching plants with holy belief (e.g. Ficus religiosa, Ocimum tenuiflorum or Ocimum sanctum) (χ2=4.22; p=0.04), participating in cultural rituals (χ2=5.20; p=0.02), consuming Prasad” "Prasad is a sacred food offering in Hinduism made during religious ceremonies, consisting of pure and natural ingredients that are distributed among devotees as a symbol of divine blessings and believed to have spiritual and medicinal benefits."(χ2=6.10; p=0.01), sleeping in usual bed (χ2=16.04; p=0.001), using packed pads (χ2=5.45; p=0.02), using common toilet (χ2=27.48; p=0.001), and using common tap (χ2=16.27; p=0.001).

However, the study did not find a significant association between perceived stress and certain restrictions, such as interacting with males (χ2=3.30; p=0.09), touching males (χ2=0.014; p=0.91), consuming meat products (χ2=0.004; 0.95), vegetables (χ2=0.007; p=0.93), fruits (χ2=0.11; p=0.74), entering inside a temple (χ2=2.41; p=0.12), going to school (χ2=1.61; p=0.20), roaming around and playing with friends (χ2=0.87; p=0.35), using heavy blankets and mattresses (χ2=2.51; p=0.11), and using sundry cloth pads (χ2=1.62; p=0.20).

### Univariate Analysis: Mentrual Restriction

**Table 4.** : Descriptive statistics of restrictions during menstrual period, n=370

| Restriction during the menstrual period | Number (%) |  |
| --- | --- | --- |
|  | Yes (%) | No (%) |
| Restricted to enter or reside inside the house | 315 (85.14) | 55 (14.86) |
| Restricted to reside inside house (Sent to Chau-Goth) | 210 (56.76) | 160 (43.24) |
| Restricted to interact with male members | 71 (19.19) | 299 (80.81) |
| Restricted to touch male members | 94 (25.41) | 276 (74.59) |
| Restricted to cook food | 318 (85.95) | 52 (14.05) |
| Restricted to touch food to be eaten by others | 322 (87.03) | 48 (12.97) |
| Restricted to eat together with family | 272 (73.51) | 98 (26.49) |
| Restricted to touch livestock or animals | 216 (58.38) | 154 (41.62) |
| Restricted to consume dairy products | 271 (73.24) | 99 (26.76) |
| Restricted to consume meat products | 53 (14.32) | 317 (85.68) |
| Restricted to consume vegetables | 52 (14.05) | 318 (85.95) |
| Restricted to consume fruits | 60 (16.22) | 310 (83.78) |
| Restricted to enter inside temple | 336 (90.81) | 34 (9.19) |
| Restricted to touch plants with holy belief | 312 (84.32) | 58 (15.68) |
| Restricted to get involve in cultural rituals of family functions | 177 (47.84) | 193 (52.16) |
| Restricted to have *Prasad* | 22 (5.95) | 348 (94.05) |
| Restricted to go to school | 38 (10.27) | 332 (89.73) |
| Restricted to roam and play around with friends | 61 (16.49) | 309 (83.51) |
| Restricted to sleep in any bed | 190 (51.35) | 180 (48.65) |
| Restricted to sleep in usual bed | 315 (85.14) | 55 (14.86) |
| Restricted to use heavy blankets and mattresses | 122 (32.97) | 248 (67.03) |
| Restricted to use packed pads | 72 (19.46) | 298 (80.54) |
| Restricted to sundry cloth pad | 74 (20.00) | 296 (80.00) |
| Restricted to use the common toilet | 153 (41.35) | 217 (58.65) |
| Restricted to use common taps | 295 (79.73) | 75 (20.27) |

Among the 370 participants included in this study, more that three quarter (85.14%) were restricted from entering or residing inside the house which is a . Over two quarters of the participants (56.67%) were sent to Chhau Goth. Although more that three quarters of adolescent girls (80.81%) were not restricted from interacting with male family members, a quarter of them (25.41%) were restricted from touching them. During menstruation, a large proportion of adolescent girls were restricted from cooking and touching food to be eaten by others, more than three quarter (85.95%) (87.03%), but slightly over a quarter (26.49%) were not restricted from eating with their family. While the majority of the participants (85.68%) were not restricted from consuming meat products, more than 2 quarters (58.38%) were restricted from touching livestock during menstruation. Additionally, almost three quarters of adolescent girls (73.24%) were restricted from consuming dairy products during menstruation. Less than one quarter reported restrictions on consuming vegetables (14.05%) and fruits (16.22%) during menstruation. Regarding religious practices, a significant proportion, almost 4 quarters of participants were restricted from entering temples (90.81%), and more than three quarters (84.32%) were restricted from touching any plants due to religious beliefs. Almost 2 quarters of the adolescent girls (47.84%) were restricted from participating in cultural rituals during menstruation, but lmost 4 quarters (94.05%) were not restricted from receiving Prasad from these rituals. A small proportion, less than a quarter of adolescent girls were restricted from attending school (10.27%) and playing with friends (16.49%) during menstruation. While slightly over two quarter of the participants (51.35%) were restricted from sleeping in any bed during menstruation, ore than three quarters (85.14%) were restricted from sleeping in their usual bed. Furthermore, more than one quarter of the participants reported restrictions on using heavy blankets and mattress (32.97%). Less than one quarter of the population were restricted to use packed or even sundry clothe pads (19.46% and 20.00%), respectively during menstruation. More than three quarters of the participants were restricted from using common taps (79.73%) and whereas almost two quarters were restricted to use common toilets (41.35%) during menstruation.

### Univariate Analysis: Perceived Stress Scale

**Table 5.**
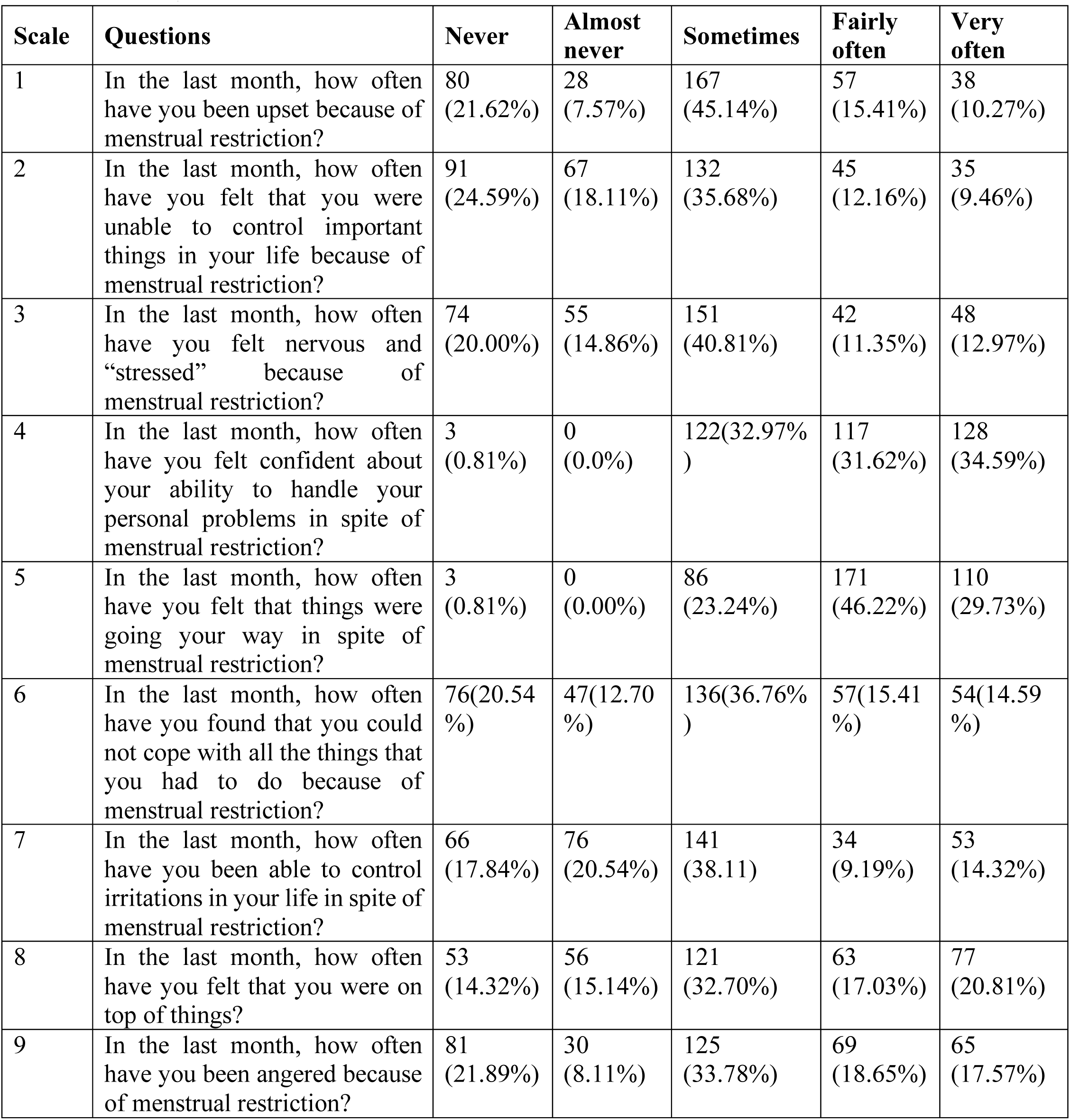
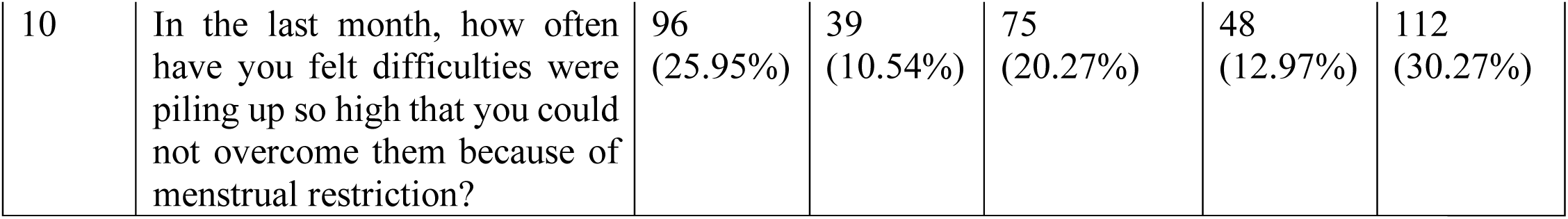
Descriptive statistics of menstrual restrctions among school going adolescent girls measured using Perceived Stress Scale, n=370

Table 5 shows the stress due to menstrual restriction measured by five point perceived stress scale shows less than one quarter (12.97%) of adolescent girls very often felt nervous and “stressed” and (10.27%) been upset because of menstrual restriction. More than one tquarter (32.70%, 32.97%) of the adolescent girls only sometimes respectively felt that they were on the top of things, and felt confident about their ability to handle personal problems. More than one quarter (30.27%) of the participants very often felt difficulties were piling up so high that they could not overcome them because of menstrual restriction. However, almost one quarter (25.94%) of the participants never felt so. Adolescent girls who sometimes been angered because of menstrual restriction were more than one quarters ( 33.78%) while less than one quarter (18.65%) of the adolescent girls been angered fair often because of menstrual restriction. Less than one quarter (20.54%) of the adolescent girls almost never been able to control irritations in their life in spite of menstrual restriction. However more than one quarter (38.11%) sometimes been able to control irritations in their life. Among all 370 adolescent girls, less than one quarter (14.59%) of them very often felt that they could not cope with all the things that they had to do because of menstrual restriction. And less than one quarter (12.16%) of them felt that they were unable to control important things in life because of menstrual restriction. Still near about two quarter (46.22%) of the adolescent girls fair often felt that things were going their way in spite of menstrual restriction.

### Severity of perceives Stress

**Table 6.** Severity of perceived stress level as per Global Measure of Perceived Stress (PSS) scale, n=370 (16)

| Perceived Stress Level | Number (%) |
| --- | --- |
| Low Stress ($\leq 20$) | 187 (50.54) |
| High Stress ($> 20$) | 183 (49.46) |

Almost two quarters (49.46%) of the respondents reported high perceived stress due to menstrual restriction (Table 5).

### Multi-Variate Analysis: Logistic regression analysis of socio-demographic variables with perceived stress level

**Table 7.** Logistic regression analysis of socio-demographic variables with perceived stress level among school going adolescent girls

| Variables | Perceived Stress Level<br>n (%) |  | cOR, 95% CI | p-value | aOR, 95% CI | p-value |
| --- | --- | --- | --- | --- | --- | --- |
|  | High | Low |  |  |  |  |
| **Age** |  |  |  |  |  |  |
| 14 | 54 (45.76) | 64 (54.24) | 0.77 (0.43-1.40) | 0.39 | 0.62 (0.30-1.23) | 0.17 |
| 15 | 44 (41.90) | 61 (58.10) | 0.66 (0.36-1.22) | 0.18 | 0.61 (0.31-1.19) | 0.15 |
| 16 | 49 (62.82) | 29 (37.18) | 1.55 (0.80-2.99) | 0.19 | 1.67 (0.84-3.34) | 0.14 |
| 17 | 36 (52.17) | 33 (47.83) | Ref |  | Ref |  |
| **Grade** |  |  |  |  |  |  |
| 8 | 78 (57.78) | 57 (42.22) | 1.51 (0.89-2.55) | 0.12 | 2.04 (1.09-3.97) | **0.024** |
| 9 | 58 (42.65) | 78 (57.35) | 0.82 (0.49-1.39) | 0.46 | 1.09 (0.62-1.90) | 0.87 |
| 10 | 47 (47.47) | 52 (52.53) | Ref |  | Ref |  |
| **Ethnicity** |  |  |  |  |  |  |
| Dalit | 100 (55.56) | 80 (44.44) | Ref |  | Ref |  |
| Janajati | 5 (13.89) | 31 (86.11) | 1.23 (0.05-0.35) | **0.001** | 2.12 (1.13-3.97) | **0.01** |
| Brahmin/C<br>hhetri | 78 (50.65) | 76 (49.35) | 0.82 (0.53-1.26) | 0.37 | 1.04 (0.58-1.85) | 0.88 |

Table 7 shows the results of logistic regression analysis of the socio-demographic variables with perceived stress level due to menstrual restrictions among adolescent girls. By adjusting the effects of other explanatory variables the odds of perceived stress level increases by 2.04 times among the adolescent girls of grade eight than those of grade ten (OR= 2.04; CI =1.09-3.97). Crude odds ratio shows Janajati adolescent girls had 1.23 times higher odds of perceived stress than Dalit adolescent girls (OR=1.23; CI=0.05-0.35). While adjusting effects of other predictors, the odds of perceived stress increases by 2.12 times among Janajati girls than Dalit adolescent girls (OR=2.12; CI=1.13- 3.97). There was no any relationship found between age group of adolescent girls and their perceived stress level due to menstrual restriction.

### Logistic regression analysis of menstrual restrictions with perceived stress

**Table 8.** Logistic regression analysis of menstrual restrictions with perceived stress level among school going adolescent girls

| Variables | Perceived Stress Level<br>n (%) |  | cOR, 95% CI | p-value |
| --- | --- | --- | --- | --- |
|  | High | Low |  |  |
| Restricted to enter or reside inside the house<br>Yes (315)<br>No (55) | 170 (53.97%)<br>13 (23.64%) | 145 (46.03%)<br>42 (76.36%) | 3.78 (1.96-7.33) | **0.001** |
| Restricted to reside inside house (Sent to Chau-Goth)<br>Yes (210)<br>No (160) | 128 (60.95%)<br>55 (34.38%) | 82 (39.05%)<br>105 (65.63%) | 2.98 (1.94-4.57) | **0.001** |
| Restricted to touch food to be eaten by others<br>Yes (322)<br>No (48) | 169 (52.48%)<br>14 (29.17) | 153 (47.52%)<br>34 (70.83%) | 2.68 (1.38-5.19) | **0.003** |
| Restricted to cook food<br>Yes (318)<br>No (52) | 169 (53.14%)<br>14 (26.92%) | 149 (46.86%)<br>38 (73.08%) | 3.08 (1.61-5.90) | **0.001** |
| Restricted to touch plants with holy belief (e.g. *Bar*, *Pipal*, *Tulsi* etc.)<br>Yes (312)<br>No (58) | 162 (51.92%)<br>21 (36.21%) | 150 (48.08%)<br>37 (63.79%) | 1.90 (1.07-3.39) | **0.03** |
| Restricted to touch livestock or animals<br>Yes (216)<br>No (154) | 130 (60.19%)<br>53 (34.42%) | 86 (39.81%)<br>101 (65.58%) | 2.88 (1.87-4.43) | **0.001** |
| Restricted to eat together with Family<br>Yes (272)<br>No (98) | 150 (55.15%)<br>33 (33.67%) | 122 (44.85%)<br>65 (66.33%) | 2.42 (1.49-3.92) | **0.001** |
| Restricted to consume dairy products<br>Yes (271)<br>No (99) | 152 (56.09%)<br>31 (31.31%) | 119 (43.91%)<br>68 (68.69%) | 2.80 (1.72-4.56) | **0.001** |
| Restricted to participate in cultural rituals<br>Yes (177)<br>No (193) | 99 (55.93%)<br>84 (43.52%) | 78 (44.07%)<br>109 (56.48%) | 1.65 (1.09-2.48) | **0.017** |
| Restricted to have *Prasad*<br>Yes (22)<br>No (348) | 17 (77.27%)<br>166 (47.70%) | 5 (22.73%)<br>182 (52.30%) | 3.73 (1.35-10.3) | **0.011** |
| Restricted to sleep in any bed<br>Yes (190)<br>No (180) | 114 (60.00%)<br>69 (38.33%) | 76 (40.00%)<br>111 (61.67%) | 3.79 (1.96-7.33) | **0.001** |
| Restricted to sleep in usual bed<br>Yes (315)<br>No (55) | 170 (53.97%)<br>13 (23.64%) | 145 (46.03%)<br>42 (76.36%) | 2.41 (1.59-3.66) | **0.001** |
| Restricted to use packed pads<br>Yes (72)<br>No (298) | 45 (62.50%)<br>138 (46.31%) | 27 (37.50%)<br>160 (53.69%) | 1.93 (1.14-3.28) | **0.015** |
| Restricted to use the common toilet<br>Yes (153)<br>No (217) | 101 (66.01%)<br>82 (37.79%) | 52 (33.99%)<br>135 (62.21%) | 3.19 (2.08-4.93) | **0.001** |
| Restricted to use common taps<br>Yes (295)<br>No (75) | 162 (54.92%)<br>21 (28.00%) | 133 (45.08%)<br>54 (72.00%) | 3.13 (1.80-5.45) | **0.001** |

When looking at the relationship of menstrual restrictions with perceived stress level, respondents with restriction to enter or reside inside the house has 3.78 times higher odds of perceived stress compared with those without restriction (OR=3.78; CI=1.96-7.33). Likewise, the odds of perceived stress increases by 2.98 times for respondents who sent to *Chhau Goth* during menstruation than who resides into their own home (OR=2.98; CI=1.94-4.57). The odds of perceived stress increases by 3.08 times for respondents who were restricted to touch food to be eaten by others than those without restriction (OR=3.08; CI=1.61-5.90). Respondents who were restricted to cook food during menstruation had 3.08 times higher odds of perceived stress compared to those without restriction (OR=3.08; CI=1.61-5.90). Similarly, the odds of perceived stress was greater by 1.90 times among respondents who were restricted to touch plants with holy belief (e.g. *Bar, Pipal, Tulsi* etc.) than those without restriction (OR=1.90; CI=1.07-3.39). Respondents who were restricted to touch livestock or animals had 2.88 times higher odds of perceived stress compared to those with such restriction (OR=2.88; CI=1.87-4.43). Regarding the restriction to consume dairy products during menstruation, the odds of perceived stress was 2.80 times higher for respondents with restriction than those without restriction (OR=2.80; CI= (1.72- 4.56). The odds of perceived stress increases by 2.42 times among the respondents who were restricted to eat together with family during menstruation compared to those without restriction (OR=2.42; CI=1.49-3.92). Respondents who were restricted to participate in cultural rituals had 1.65 times higher odds of perceived stress than those with no such restriction (OR=1.65; CI=1.09- 2.48). The odds of perceived stress increases by 3.73 times among respondents who were restricted to have *Prasad* during menstruation compared to those with no such restriction (OR= 3.73; CI=1.35-10.3). The odds of perceived stress was 3.79 times higher for respondents who were restricted to sleep in any bed (OR=3.79; CI=1.96-7.33) and 2.41 times higher for those with restriction to sleep in their usual bed 2.41 (1.59-3.66) compared to those with no any such restrictions. The odds of perceived stress increases by 1.93 times among the respondents who were restricted to use packed pads during menstruation than those with no such restriction (OR=1.93; CI=1.14-3.28). Respondents who were restricted to use the common toilet during menstruation had 3.19 times higher odds of perceived stress 3.19 (2.08-4.93), while perceived stress increases by 3.13 times among those who were restricted to use common taps during menstruation compared to those with no any such restrictions.

## Discussion

A descriptive analytical cross-sectional study design was used to assess the prevalence of various kind of menstrual restrictions and their association with perceived stress and the level of perceived stress among 370 adolescent girls from Far-Western Region, Kailali, Nepal in the duration of 1 month among the participants.

The result of the study indicated more than two quarters (56.76% ) of participant being abolished in the Chau-goth whereas 85.14% were restricted to enter or reside inside the home other than their own room. There are two doors to enter the houses and the menstruating girls are allowed to only enter through their designated doors during menstruation restricting mobility in other parts of the houses. People in those areas believe to have their own temple inside their houses where the believe their ancestral gods reside. The belief is that, because most homes have a prayer room, God inhabits the home.(17) Different traditional practice followed restriction to cook food (85.95%), to touch food to be eaten by others (87.95%), eat together with family (73.51%), consume dairy products (73.24%), restricted to enter inside temple (90.81%), restricted to touch plants with holy belief (84.32%), restricted to sleep in usual bed (85.14%), restricted to use common taps (79.73%) were some of the restrictions with more than half quarter of prevalence. In A community based mixed method, quantitative, cross sectional study in Pyuthan district of mid-western Nepal it was found that Most (94.5%) of them experienced Chaupadi (Menstrual Shed) during their menarche. The phenomenological approach found that Chaupadi was common. They had various infections and ill health. Mother groups were advocating to eliminate Chaupadi in their locality. (18)

This study found the prevalence of other forms of social restrictions as well with 16.49% being restricted to play around with friends, 10.27% restricted to go to school, 41.35% restricted to use the common toilet, 47.84% being restricted to get involved in cultural rituals or family functions. In the same study, 40.4% used sanitary pad during their menstrual flow. A cross-sectional study conducted in educational institutions from a major city in South India, found that socioeconomic Status (SES) of the selected girls and their age influenced choice of napkin/pad (19) which was similar to our findings with 82.7% girls saying that they were not restricted to use sanitary pads during their menstruation. While findings form the same study showed that in Rural West Bengal, 11.25% girls used sanitary pads during menstruation, and 42.5% girls used old cloth pieces 40% girls used both cloth pieces and sanitary pads during menstruation. (19) Theses finding had similar acceptance of rate for the use of sanitary pad as to ours being only 13.78% were restricted to use sanitary pads. However, there is no data saying if the restriction came from family for these cases. The living spaces during Chaupadi lacked basic necessities(20) Also finding that the participants of the study experienced several physical and psychological problems(20). Our study showed such psychological distress being highly prevalent among adolescent girls with 49.46% suggesting that they suffering from High Perceived Stress (>20), 50.54% suffering from Low Stress (<20) of Stress. With age the intensity and perception of stress seems to be increasing as well.The degree of stress experienced and the ways in which a person reacts to it can be influenced by a various number of factors such as personal characteristics, lifestyle, social support, and appraisal of the stressor(s), life events, socio-demographic and occupational variables. In the study, it was found that with age the level of perceived stress was higher. 45.76%, 41.90%, 62.82% and 52.17% for age 14, 15, 16 and 17 respectively. Among different ethnic groups present perceived stress was found to be comparatively high among Upper caste groups, Brahmin/Chhetri (50.65%) and Dalits (55.56%) since they religiously follow menstrual restriction in contrast to disadvantaged Janajati of hills International studies showed that the perception of psychological stress in the adolescent population tends to be higher among girls,regardless of age, in contrast this study showed low level of stress among adolescents which may be due to increasing maturity among adolescents with age.In Korea,the prevalence of perceived stress was observed in 30.5% of female participants. In England, of the adolescents who perceived themselves with high level of stress, 54.5% were female. In Brazil, the studies conducted with adolescents are leading to the same direction as international investigations, with prevalence of 30.1% among girls. In Rio Grande do Sul, among the adolescents who perceived themselves as stressed, 61.5% were female. (21) The results of current research suggested that in a developing society with high prevalence of stress, interventions targeted toward promoting financial and social equalities, social skills training, and healthy lifestyle may have the potential benefits for large parts of the population, most notably female and lower educated people. (22)

In addition, finding, winter in the hilly areas of Nepal, such as Achham, is harsh; yet in our observation of the living spaces during Chaupadi, we found that many girls lacked a mattress and warm blankets and rather slept on a rug or bare floor with sacks as cover. This may explain why hypothermia is a substantial problem among those practicing Chaupadi. (20) In our study 32.97% were restricted to use heavy blankets and mattresses during their menstrual period.Their findings of social restrictions while practicing Chaupadi, in terms of certain food andother daily activities, was also not surprising, participants were restricted from eating dairy products. (20) The researchers were impressed to see that most of our participants were permitted to attend school and read books while menstruating. In the past, menstruating girls were expected to halt school attendance, since one Hindu belief is that ‘Sarashwoti,’ the goddess of education, will become angry if a girl or woman reads, writes or touches books during her menstrual cycle. (20) Agreeably, 89.73% of our participants were not restricted to go to school either with 9.46% being restricted for the same.

The results of this study using logistic regression analysis of the socio-demographic variables with perceived stress level due to menstrual restrictions among adolescent girls. By adjusting the effects of other explanatory variables the odds of perceived stress level increases by 2.04 times among the adolescent girls of grade eight than those of grade ten (OR= 2.04; CI =1.09-3.97). Crude odds ratio shows Janajati adolescent girls had 1.23 times higher odds of perceived stress than Dalit adolescent girls (OR=1.23; CI=0.05-0.35). While adjusting effects of other predictors, the odds of perceived stress increases by 2.12 times among Janajati girls than Dalit adolescent girls (OR=2.12; CI=1.13- 3.97). There was no any relationship found between age group of adolescent girls and their perceived stress level due to menstrual restriction. There has been various studies showing various types of menstrual restriction through qualitative analysis and mentioning some form of psychological distress but none has been able to provide it with the quanitifiable numbers. A qualitative study done in Far-west Nepal found that, all study participants practiced the taboo of untouchability during menstruation and the majority of the girls (n = 77, 72%) practiced exile, or Chaupadi. It also suggested that the adolescent girls in thestudy faced many physical, psychological, and social problems and were restricted in terms of certain foods and other daily activities. (20) When looking at the relationship of menstrual restrictions with perceived stress level, respondents with restriction to enter or reside inside the house has 3.78 times higher odds of perceived stress compared with those without restriction (OR=3.78; CI=1.96-7.33). Likewise, the odds of perceived stress increases by 2.98 times for respondents who sent to *Chhau Goth* during menstruation than who resides into their own home (OR=2.98; CI=1.94-4.57). The odds of perceived stress increases by 3.08 times for respondents who were restricted to touch food to be eaten by others than those without restriction (OR=3.08; CI=1.61-5.90). Respondents who were restricted to cook food during menstruation had 3.08 times higher odds of perceived stress compared to those without restriction (OR=3.08; CI=1.61-5.90). Similarly, the odds of perceived stress was greater by 1.90 times among respondents who were restricted to touch plants with holy belief (e.g. *Bar, Pipal, Tulsi* etc.) than those without restriction (OR=1.90; CI=1.07-3.39). Respondents who were restricted to touch livestock or animals had 2.88 times higher odds of perceived stress compared to those without such restriction (OR=2.88; CI=1.87-4.43). Regarding the restriction to consume dairy products during menstruation, the odds of perceived stress was 2.80 times higher for respondents with restriction than those without restriction (OR=2.80; CI= (1.72-4.56). The odds of perceived stress increased by 2.42 times among the respondents who were restricted to eat together with family during menstruation compared to those without restriction (OR=2.42; CI=1.49-3.92). Respondents who were restricted to participate in cultural rituals had 1.65 times higher odds of perceived stress than those with no such restriction (OR=1.65; CI=1.09- 2.48). The odds of perceived stress increases by 3.73 times among respondents who were restricted to have *Prasad* during menstruation compared to those with no such restriction (OR= 3.73; CI=1.35-10.3). The odds of perceived stress was 3.79 times higher for respondents who were restricted to sleep in any bed (OR=3.79; CI=1.96-7.33) and 2.41 times higher for those with restriction to sleep in their usual bed 2.41 (1.59-3.66) compared to those with no any such restrictions. The odds of perceived stress increases by 1.93 times among the respondents who were restricted to use packed pads during menstruation than those with no such restriction (OR=1.93; CI=1.14-3.28). Respondents who were restricted to use the common toilet during menstruation had 3.19 times higher odds of perceived stress 3.19 (2.08-4.93), while perceived stress increases by 3.13 times among those who were restricted to use common taps during menstruation 3.13 j(1.80-5.45) compared to those with no any such restrictions among adolescent girls of Godawari Municipality, Kailali District.

Adolescent age being highly crucial phase of life is difficult for those born in under developed countries since topics of sexual and reproductive health are yet to be freely spoken and normalized. With such taboos going on regarding restrictions during menstruation can increase their risk to perceived stress which has significant association with other mental hazards. Since this population is in school, it would be extremely important that educational public policies could consider the possibility of inserting psychologists and nutritionists in the school context, in the sense of working directly with problems related with stress associated with menstrual restriction. If it is not possible to respond to this suggestion, we reinforce the need for teachers of Physical Education, Mental Health and Nutrition in their classes focusing on health education, to approach the theme using videos, seminars and scientific lectures addressed to the importance of avoiding such harmful practice and to reduce the levels of stress. For that reason, to mitigate such hazards extension of advocacy on normalizing m around menstruation, sexual and reproductive health issues should be in community level, also sharing of the experiences and proper mental health counseling should be done specified to the adolescent age group with maximum support. Dreadful practices such as Chaupadi must be very punishable and all the discriminatory practices should be ruled out from its root to prevent further consequences in young girl’s psychosocial well-being and life.For further research this type of study can be conducted among the larger population in community and institutional basis. View of men shall also be explored and factors associated with Chaupadi practice and other forms of menstrual restrictions can be studied as well.

One of the strong points of the study is the use of a random sample, representative of the adolescents; besides, it is one of the first representative studies relating to Menstrual restriction, looking into broader aspect of mental health of young, adolescent girls especially to perceived stress.

Since, due to being a undergraduate led research with time and budget constraints the representative sample size may be more previledged than those on remote hills of Far-Western Region as this part of Godawari Municipality was Terai region. But it also means the status in hilly regions can be significantly more risky than this. Recall bias may have impacted some of the responses.Absentee students or girls who were never enrolled into schools were also not included in the study. In the furute, a prospective cohort study with a larger sample size is needed to provide more rigorous evidence on Menstrual restriction and perceived Stress young adolescent girls. The inclusion of girls aged 14-17 is also one of its limitations.

## Data Availability

I have data available in SPSS which I can supply on request.

## Other information

### Annex 1 Questionnaires

#### Section: A Socio-demographic information

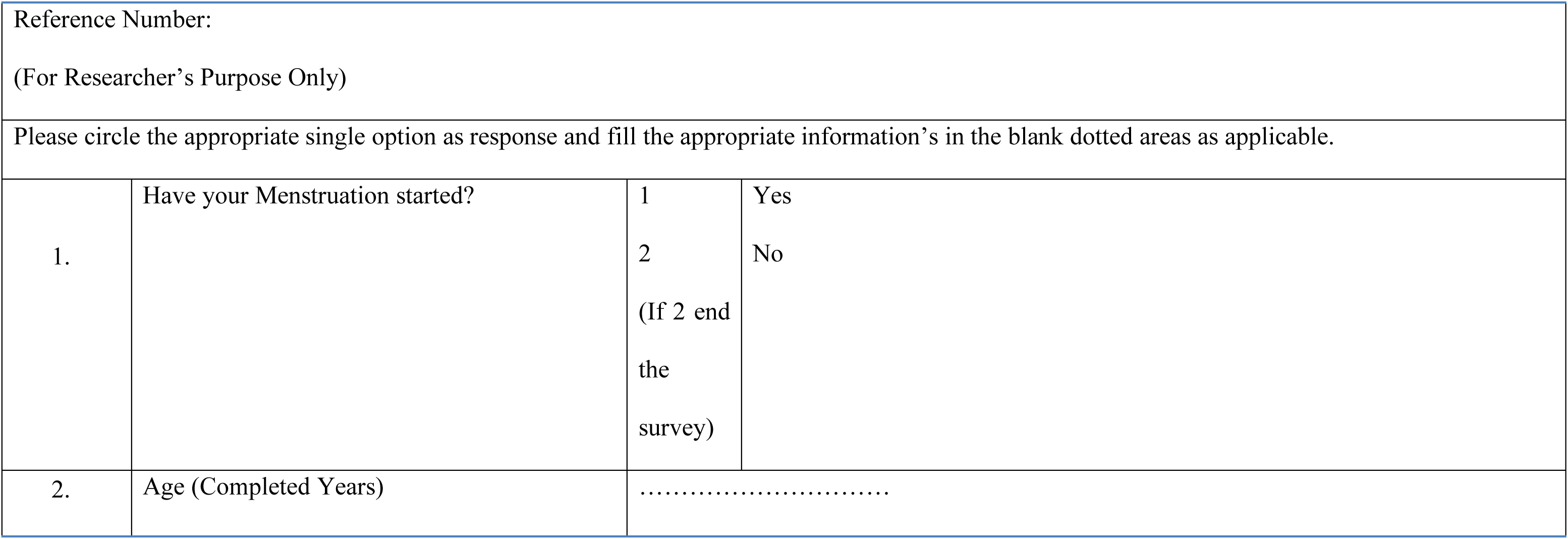
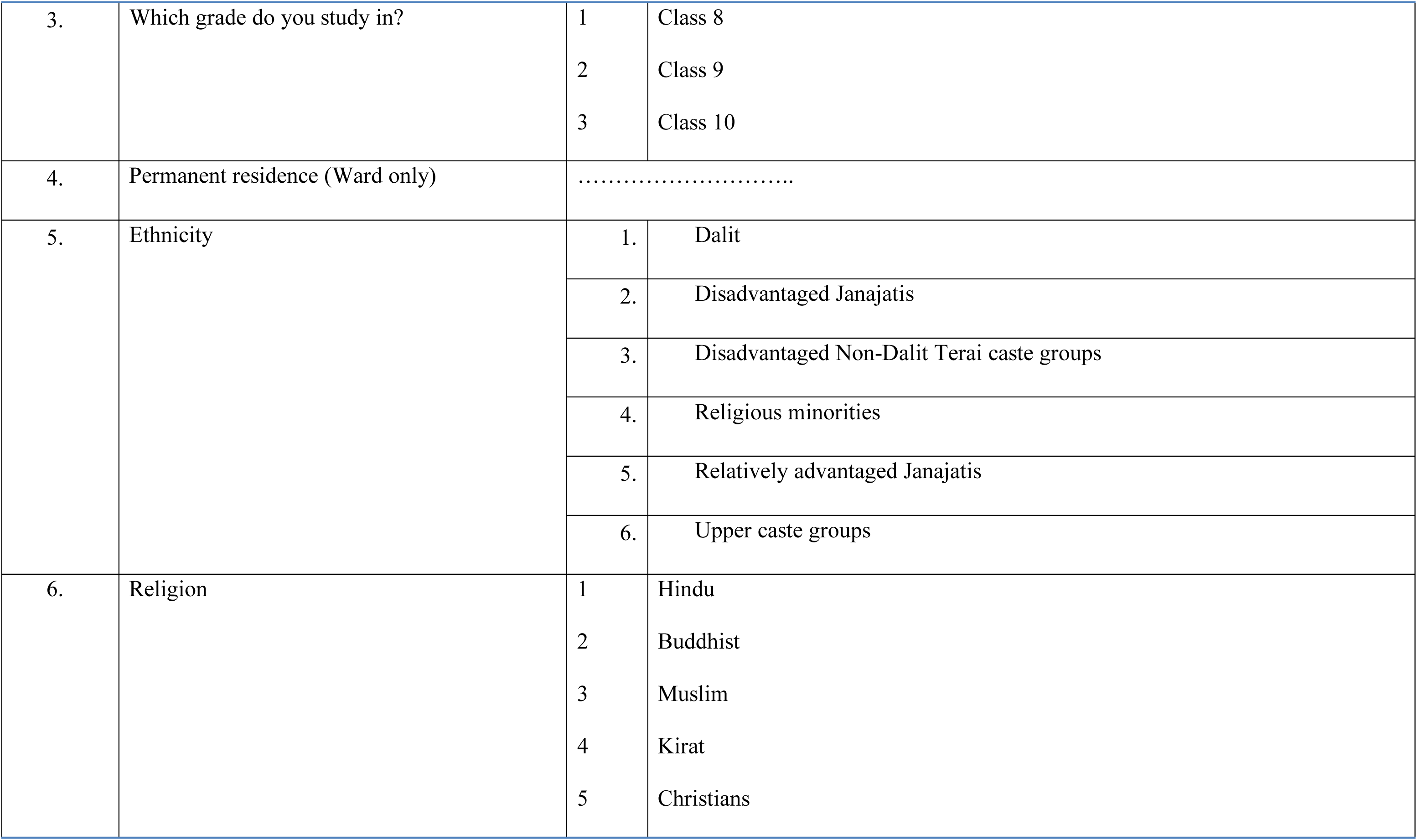
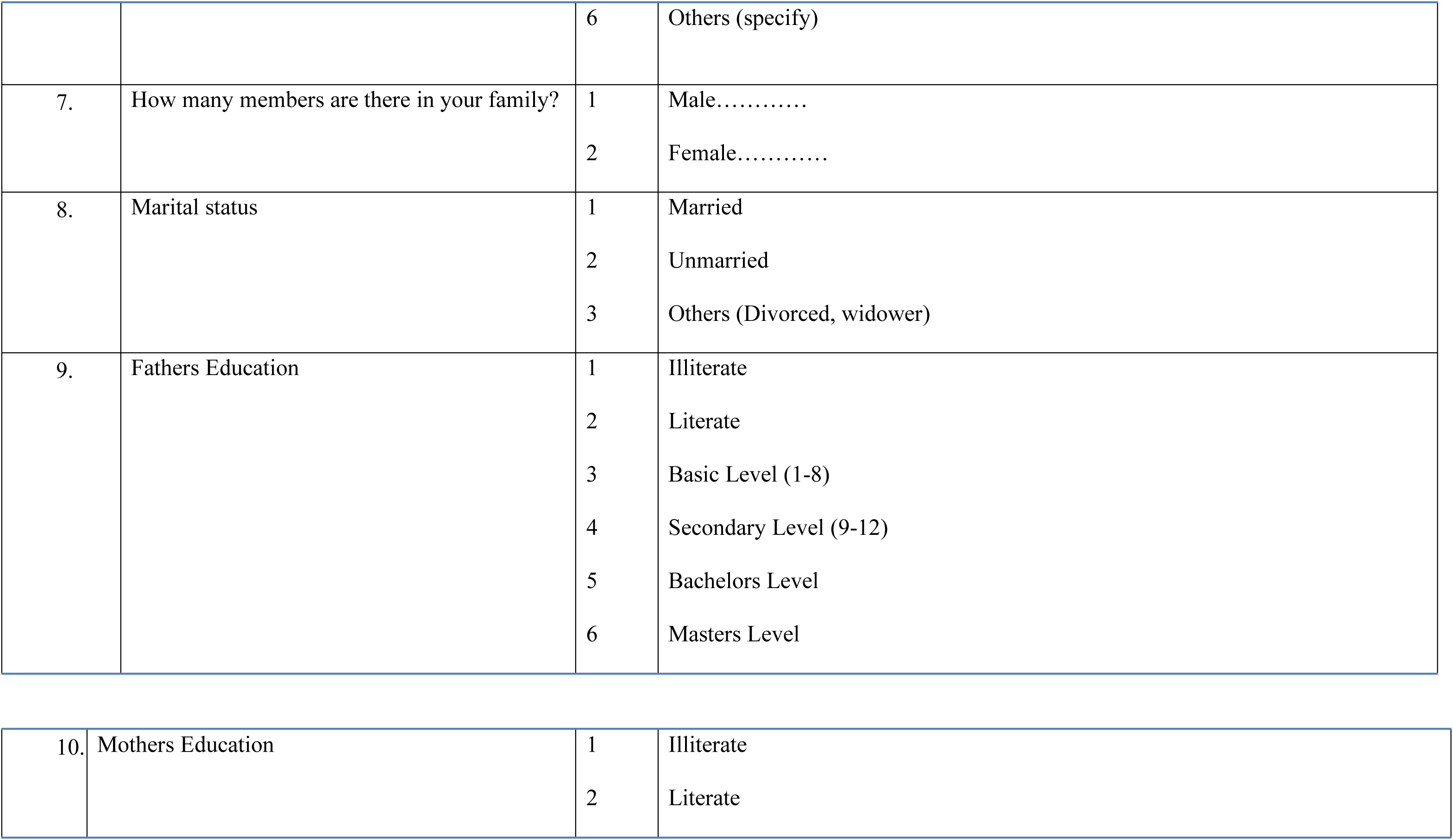
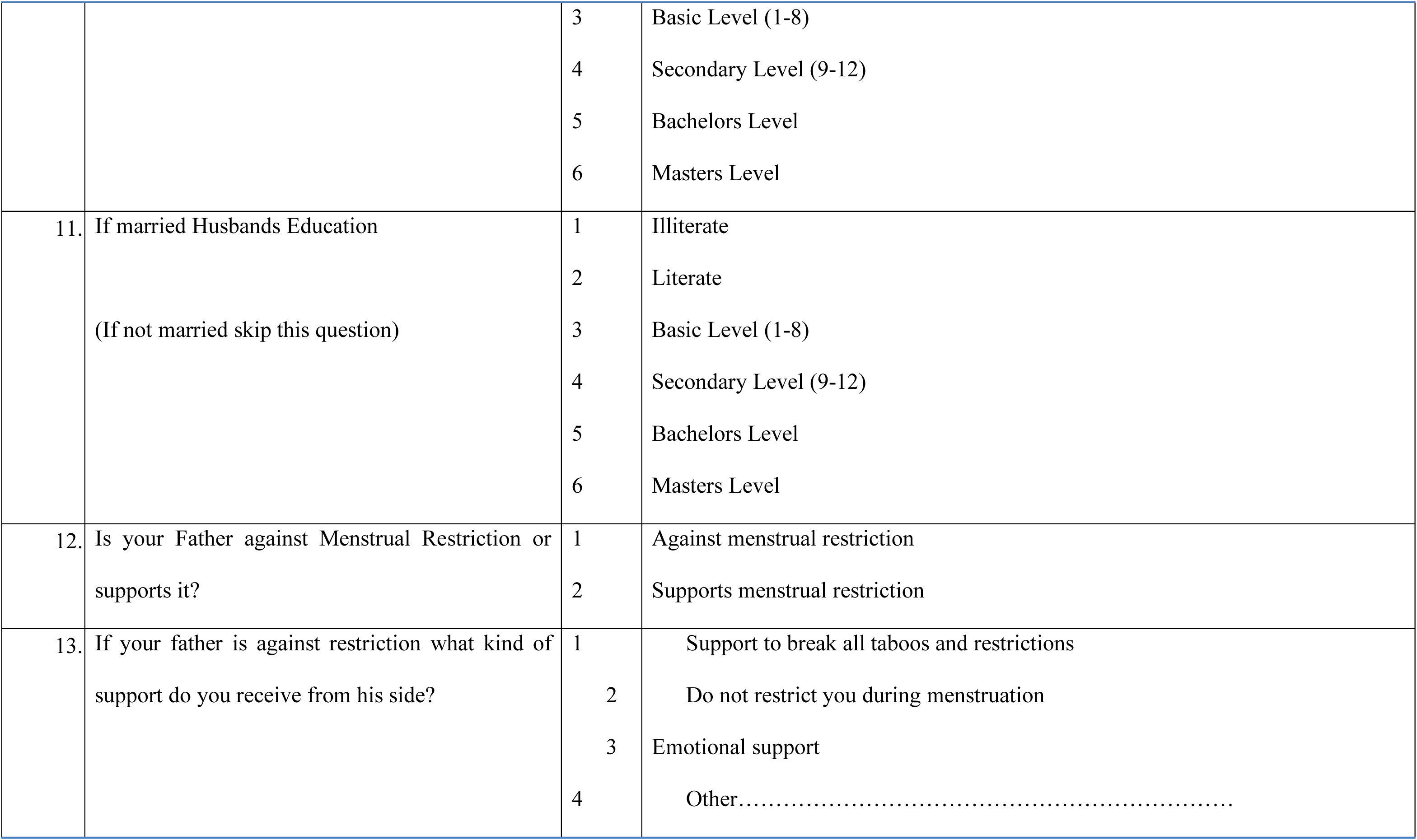
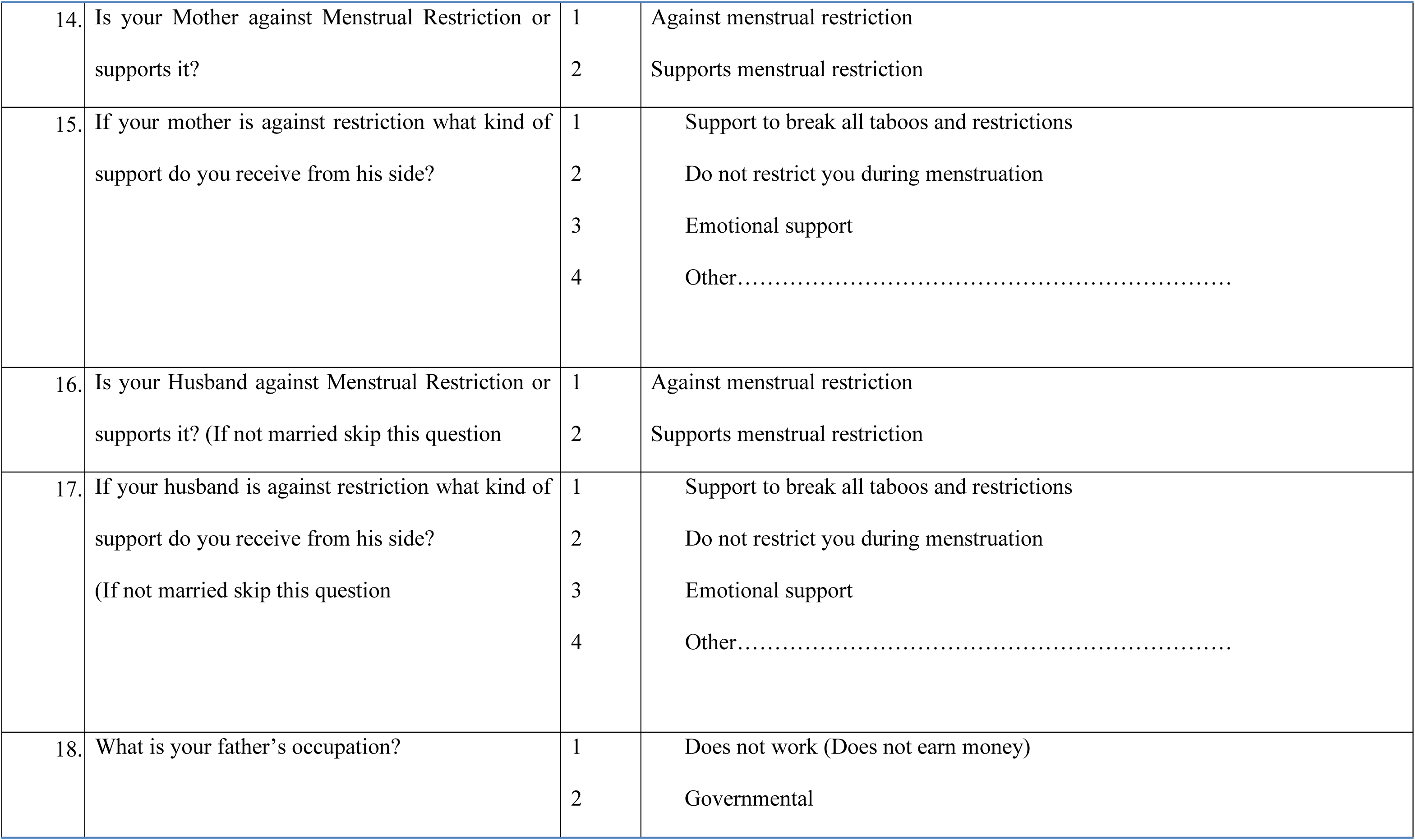
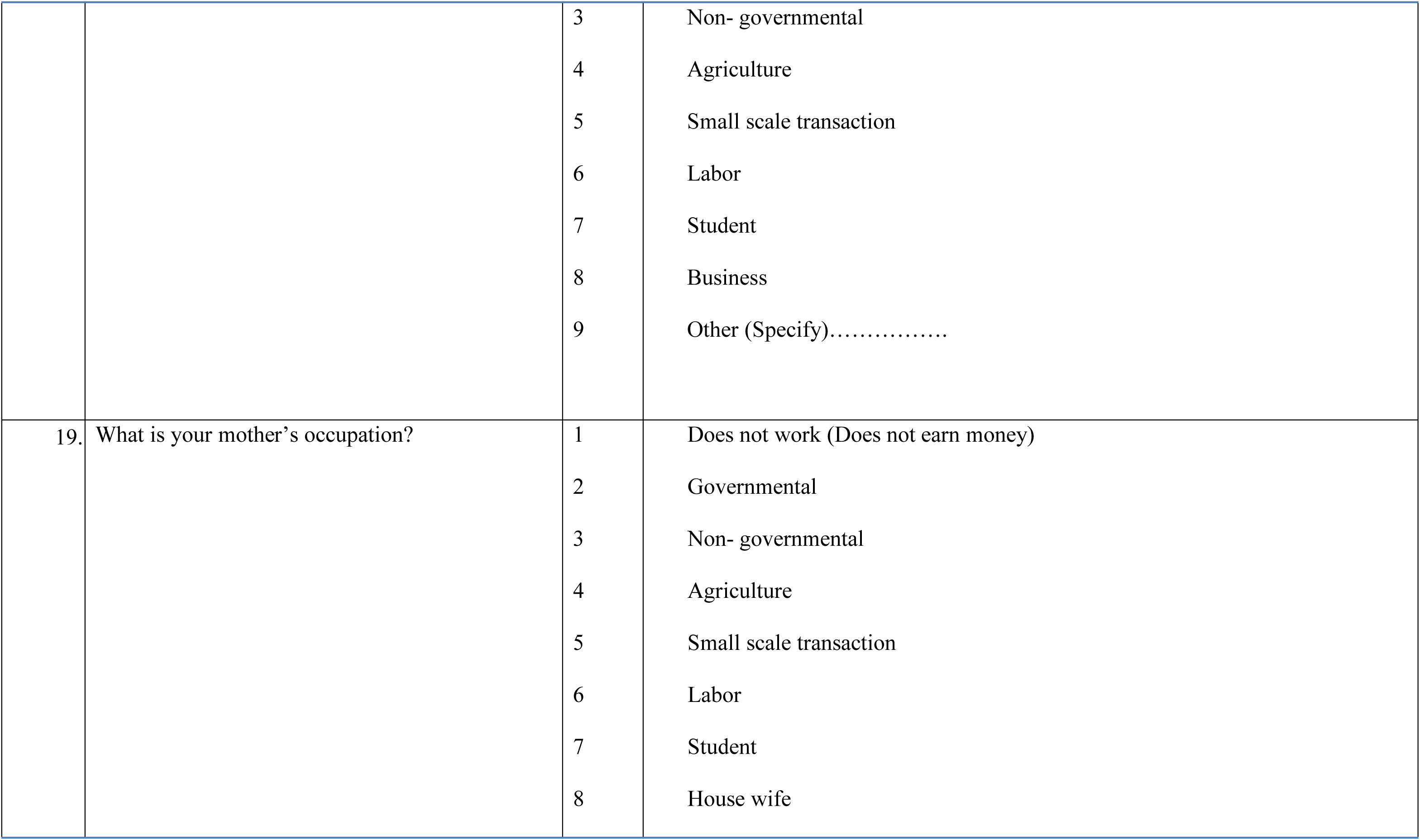
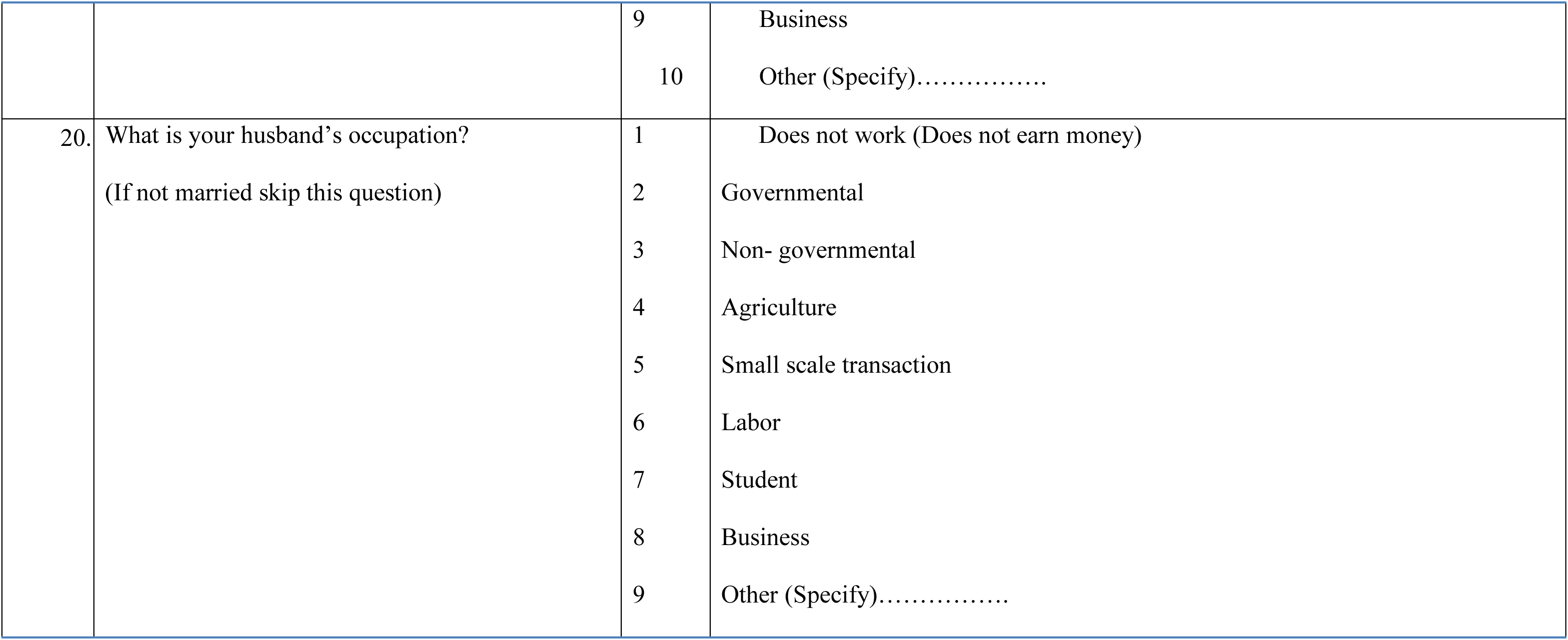

#### Section B: Menstrual Restriction

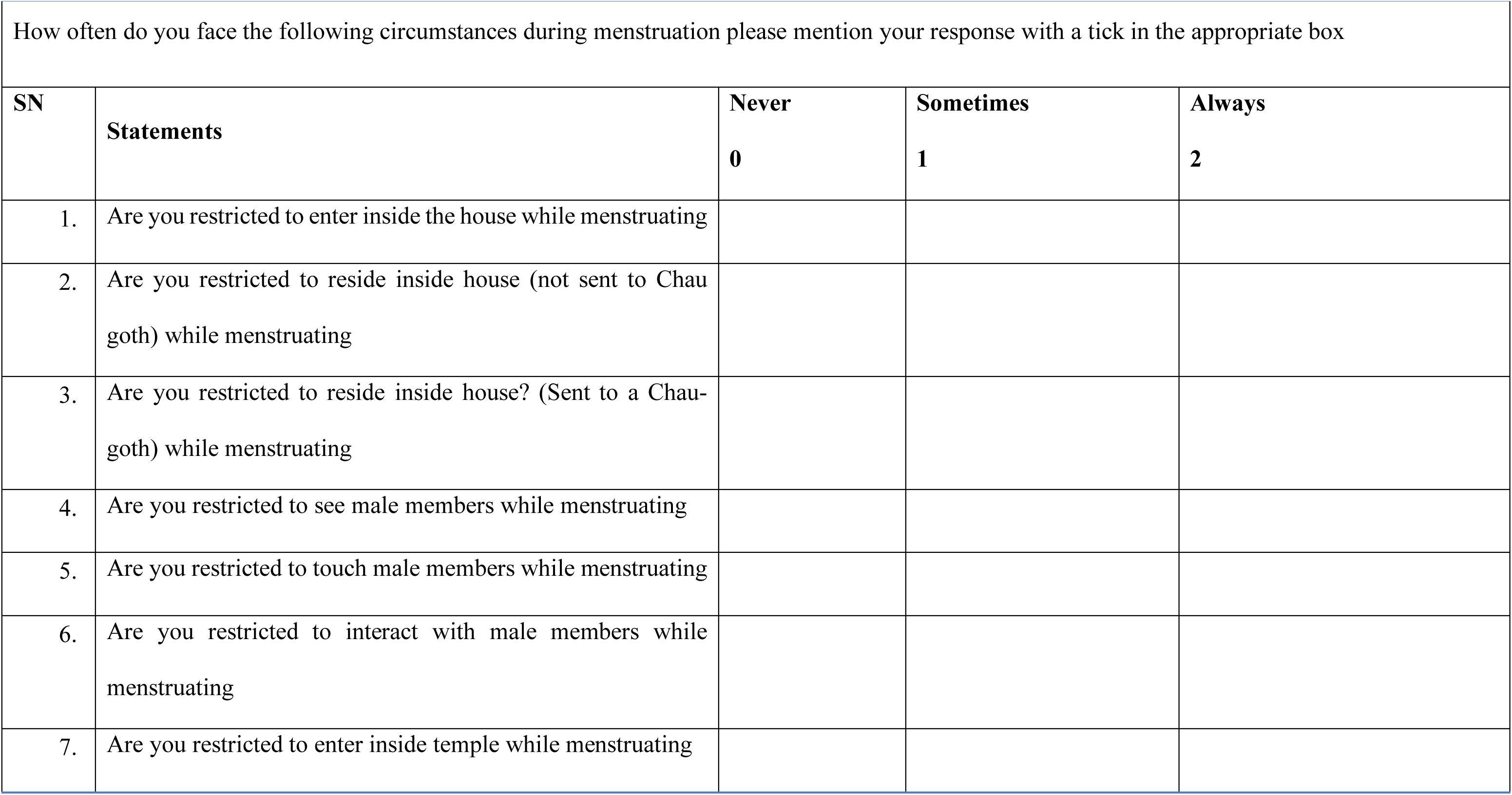
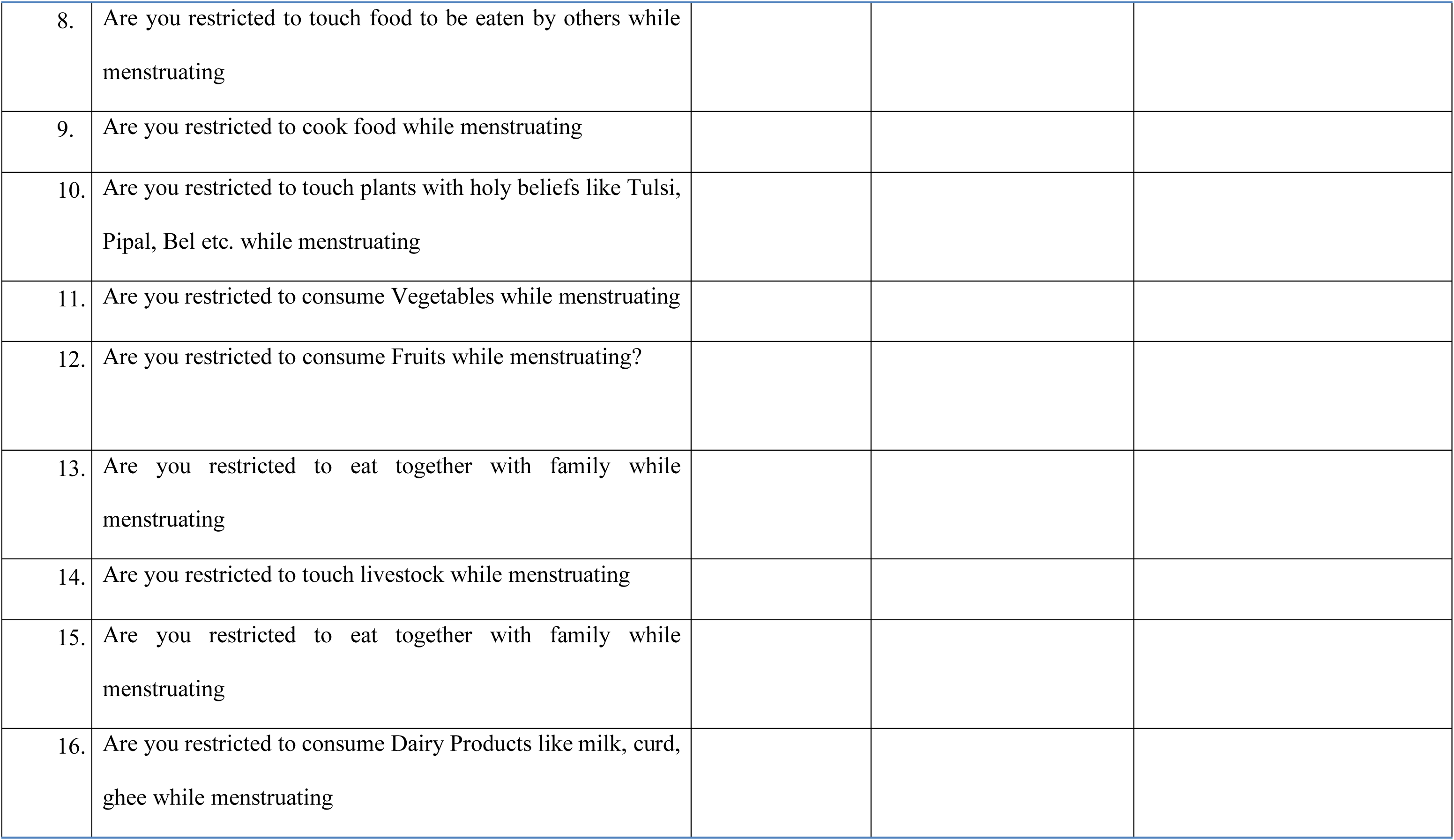
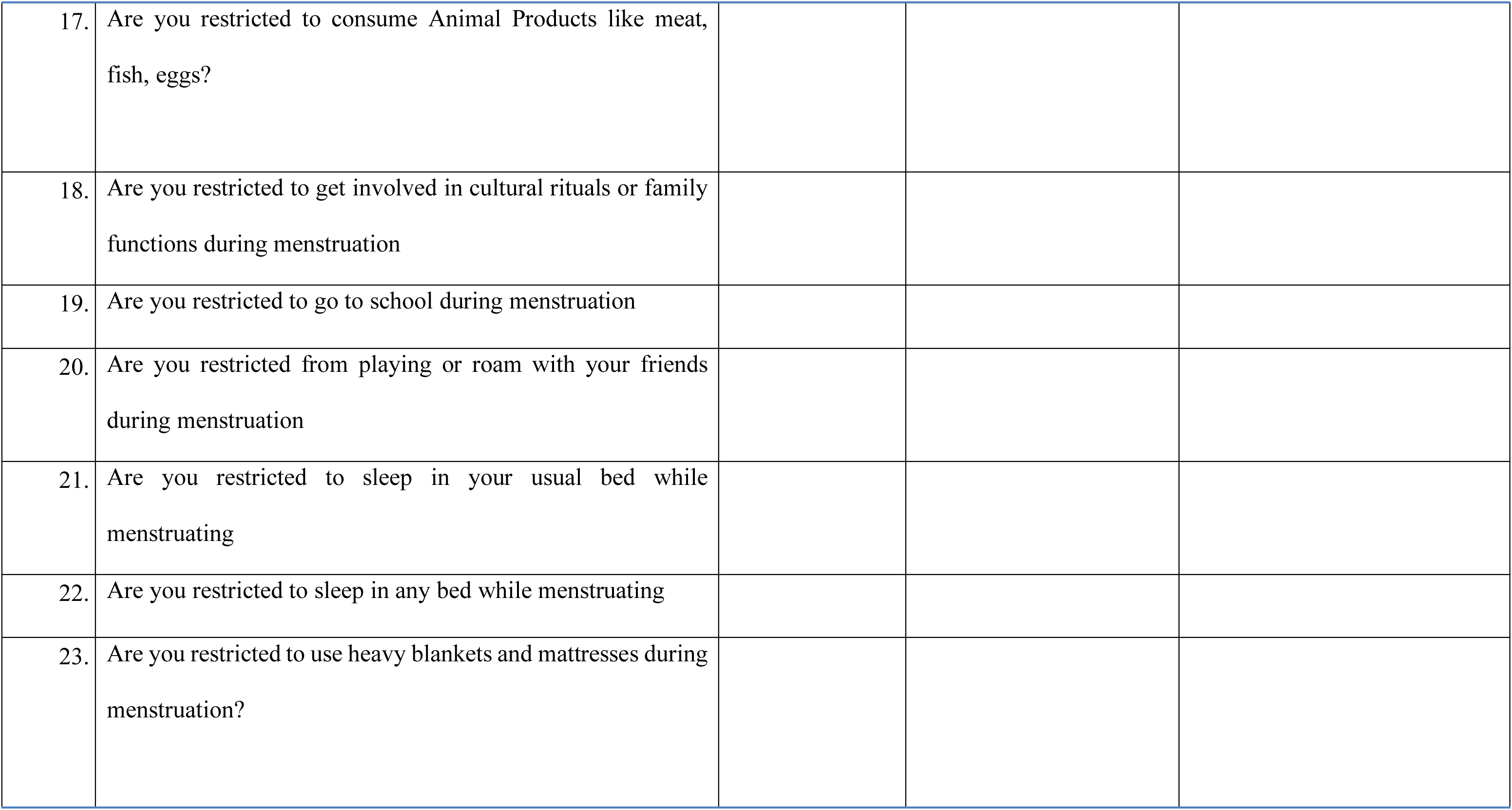
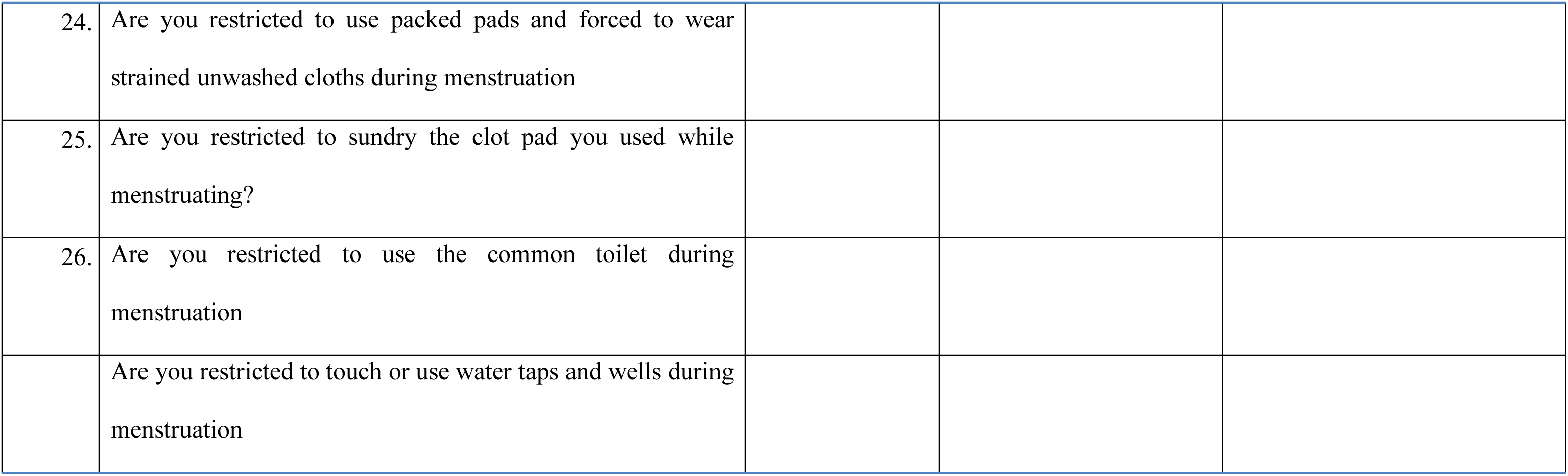

#### Section C: Perceived Stress

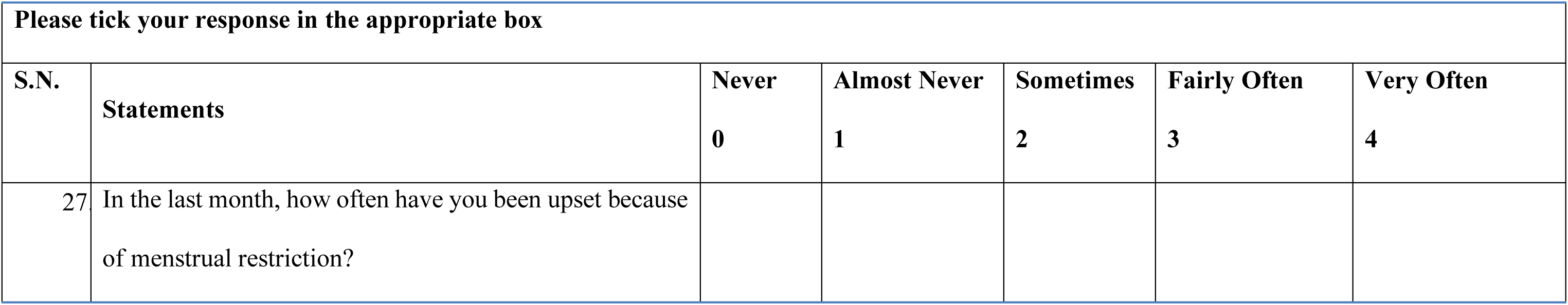
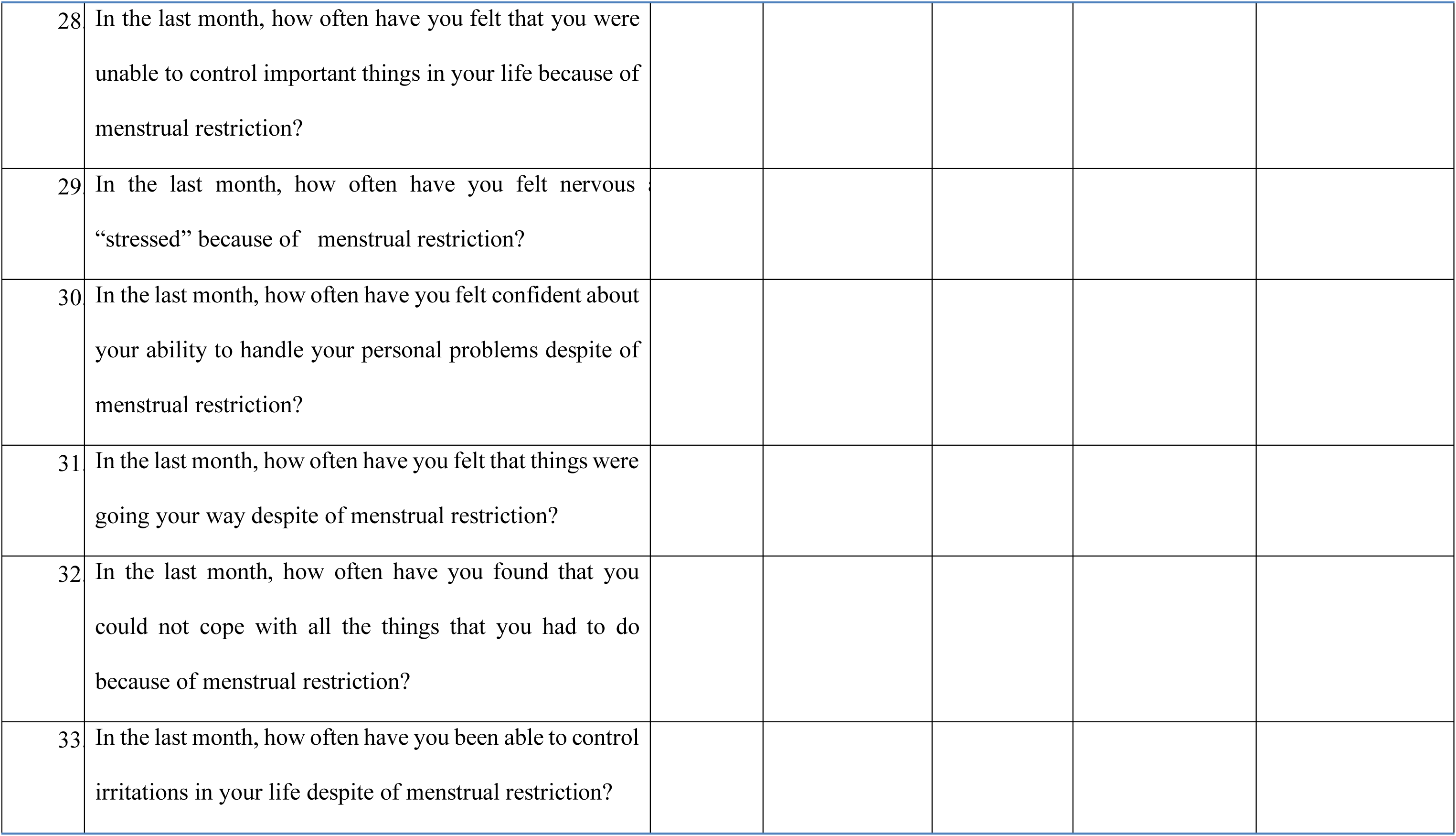
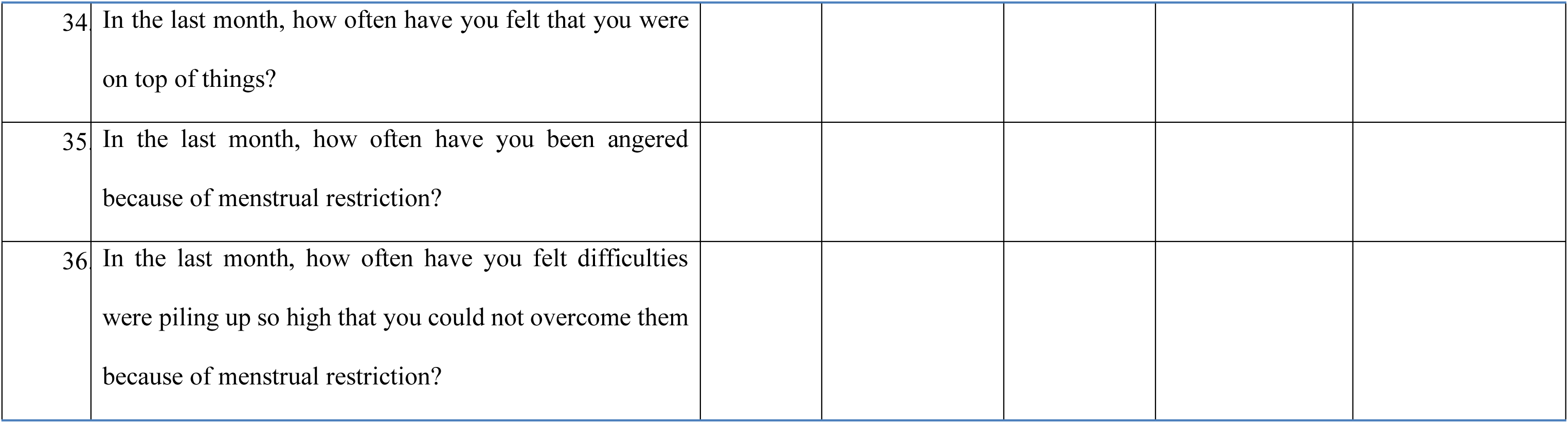

**Thank You for your Support.**

### Annex 2 Conscent Forms (Parents)

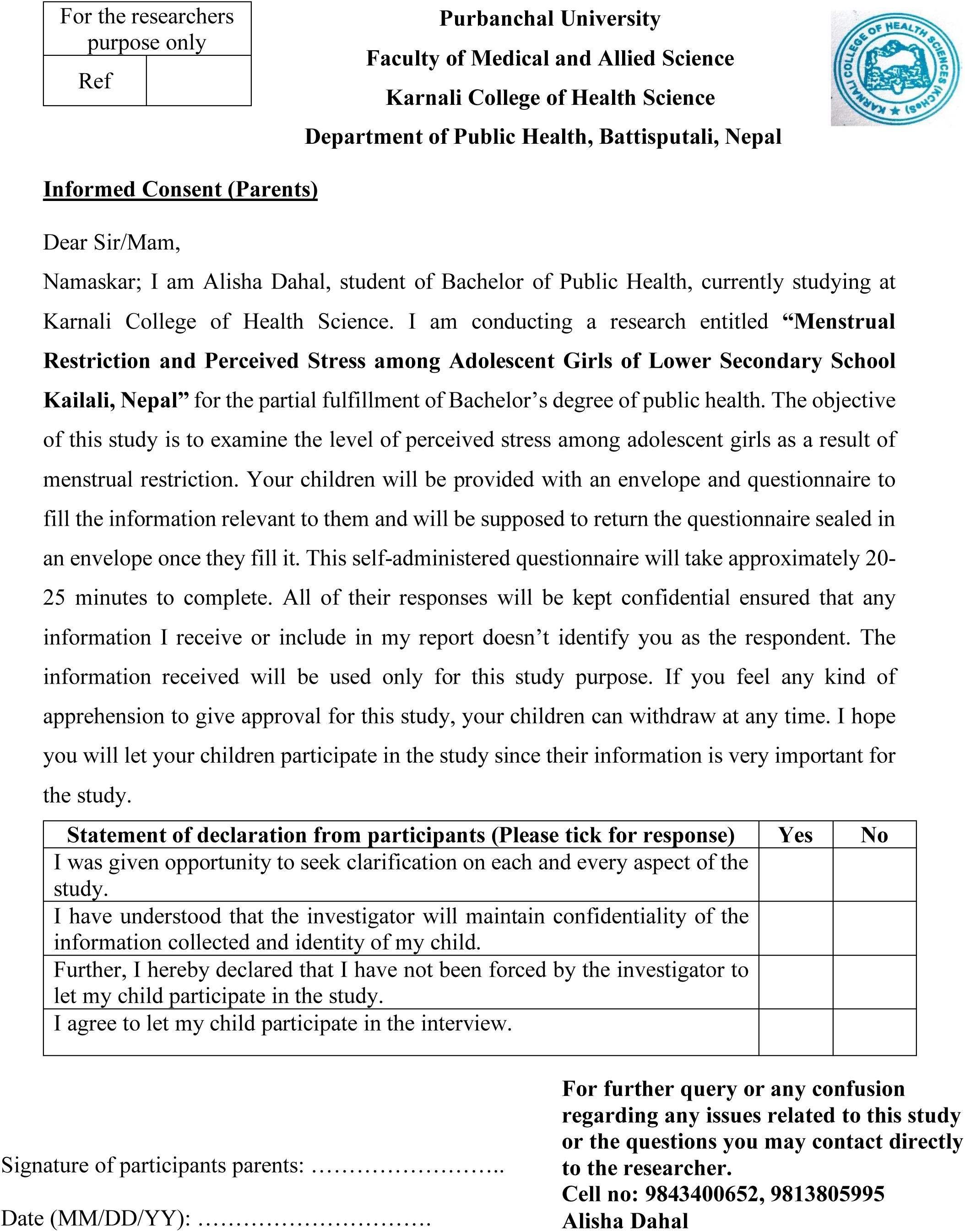

### Annex 3 Conscent Forms (Children)

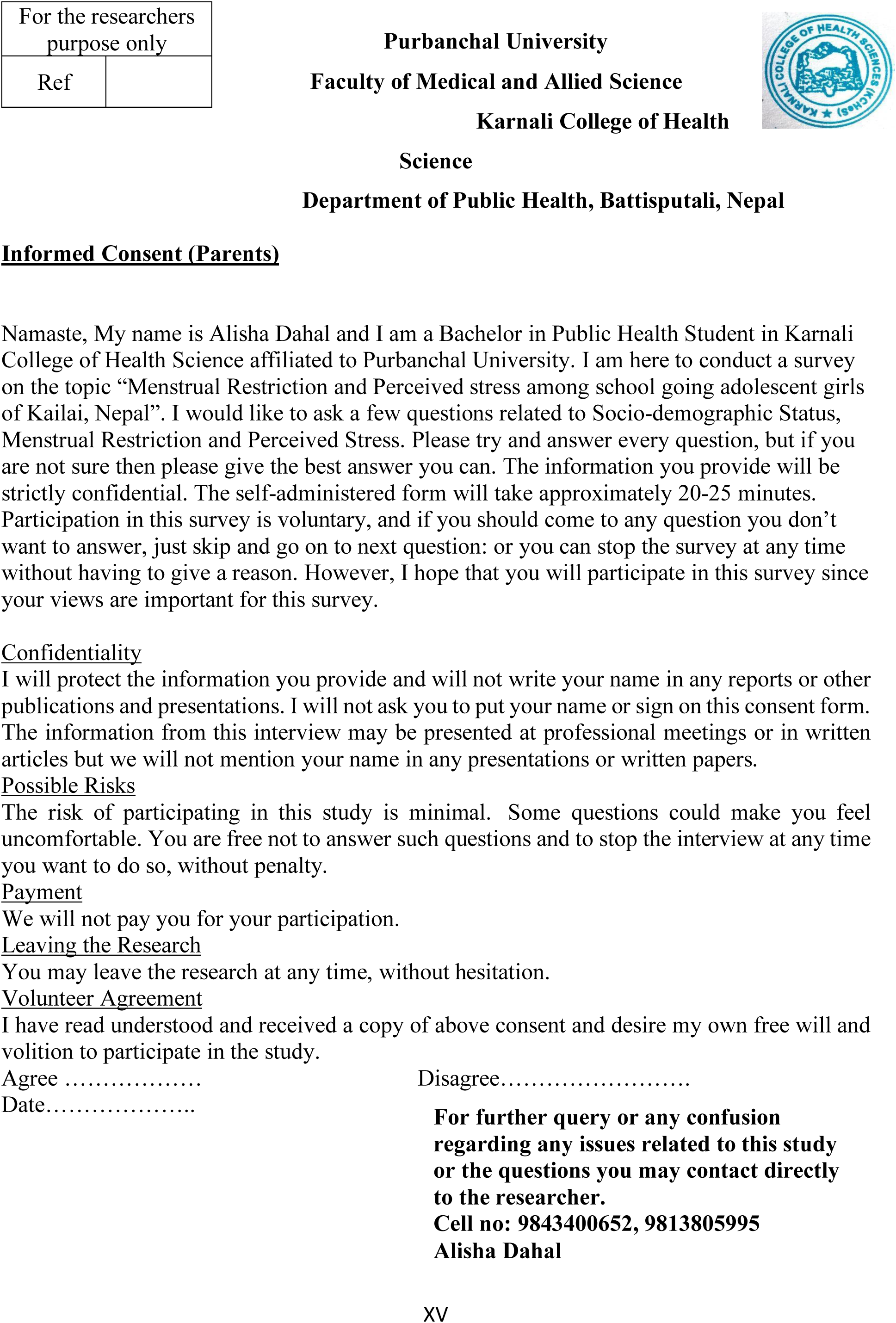

### Annex 4 PLOS ONE Clinical Studies Checklist

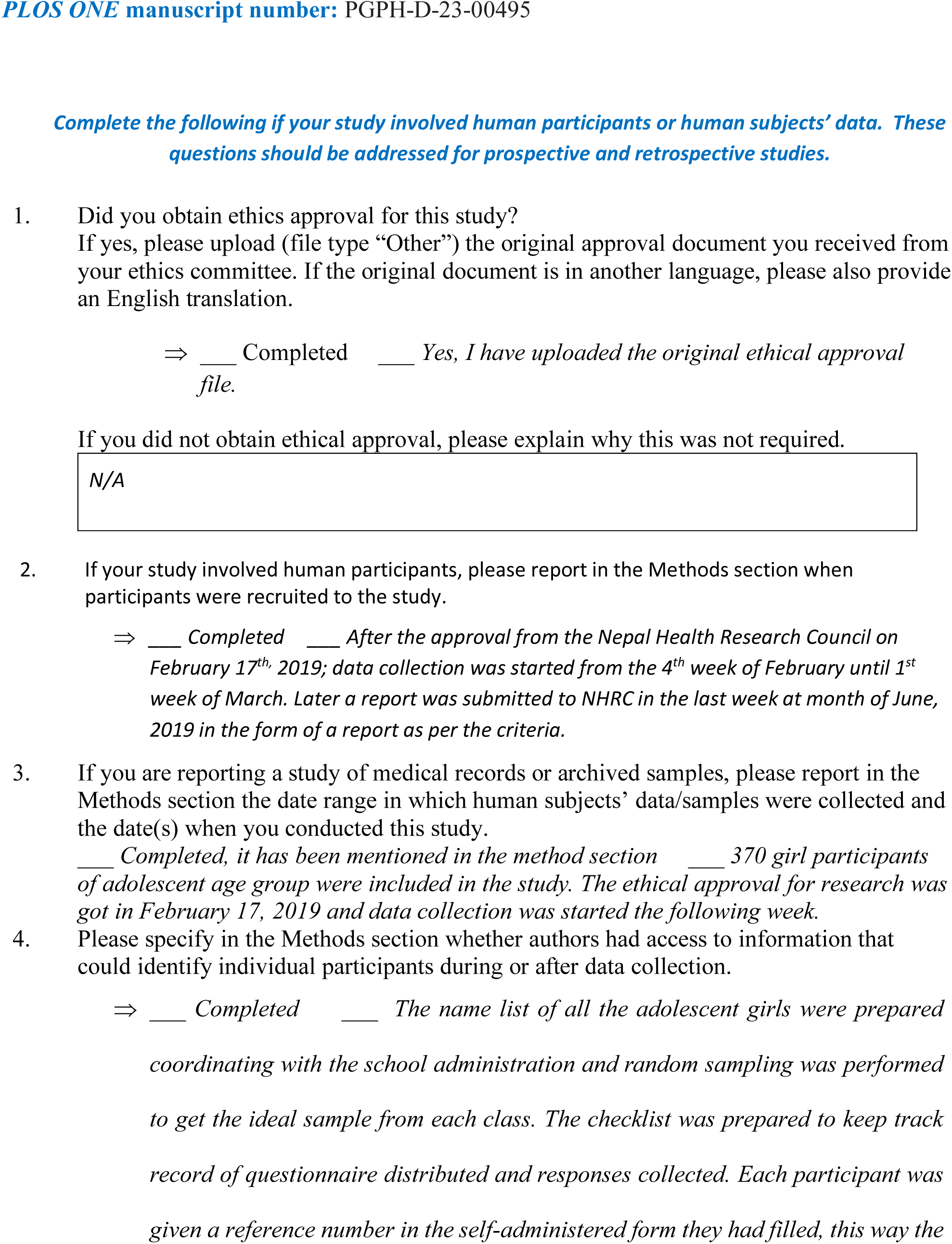
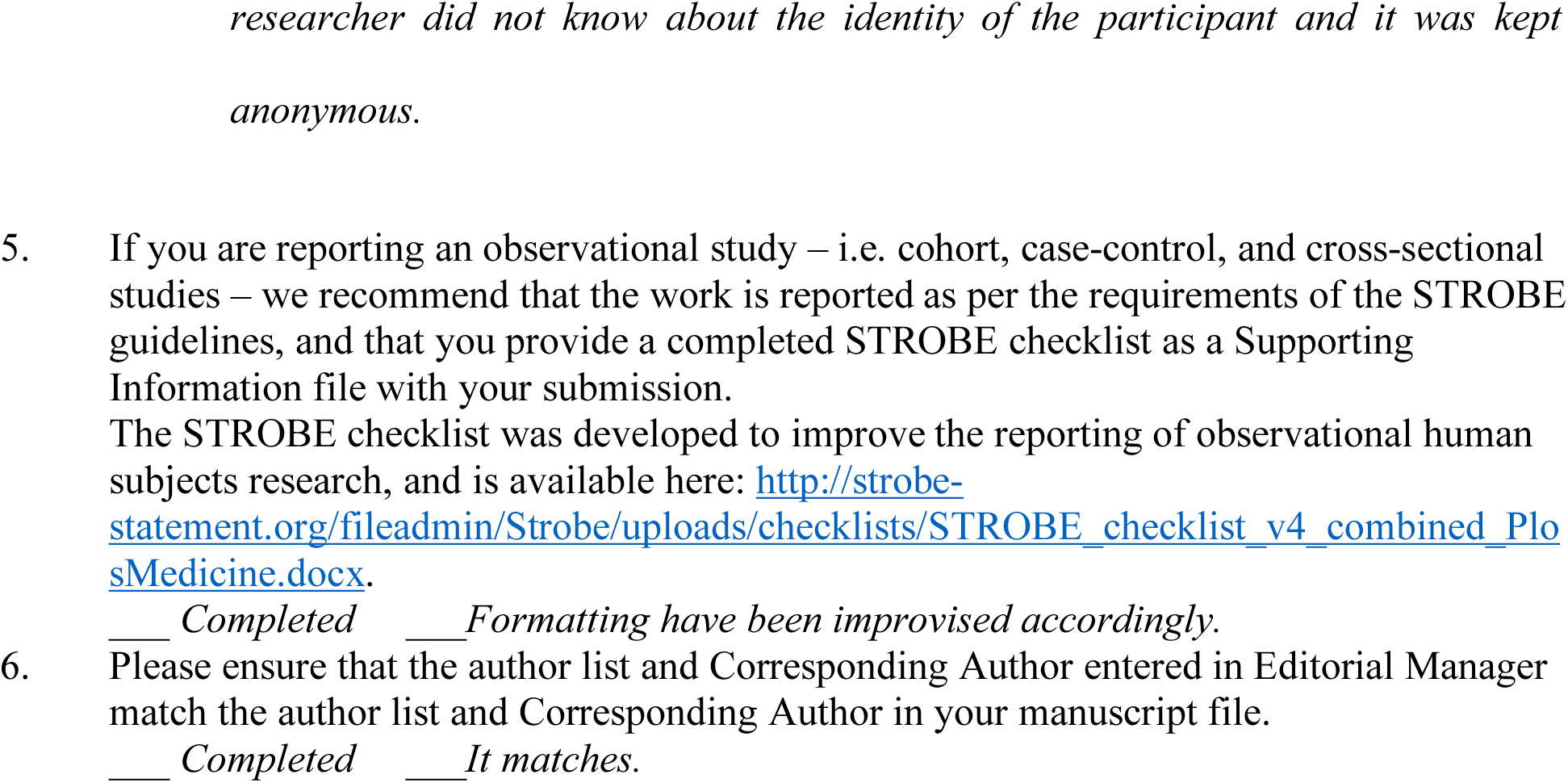

### Annex 5 NHRC Acknowledgement of the Final Report

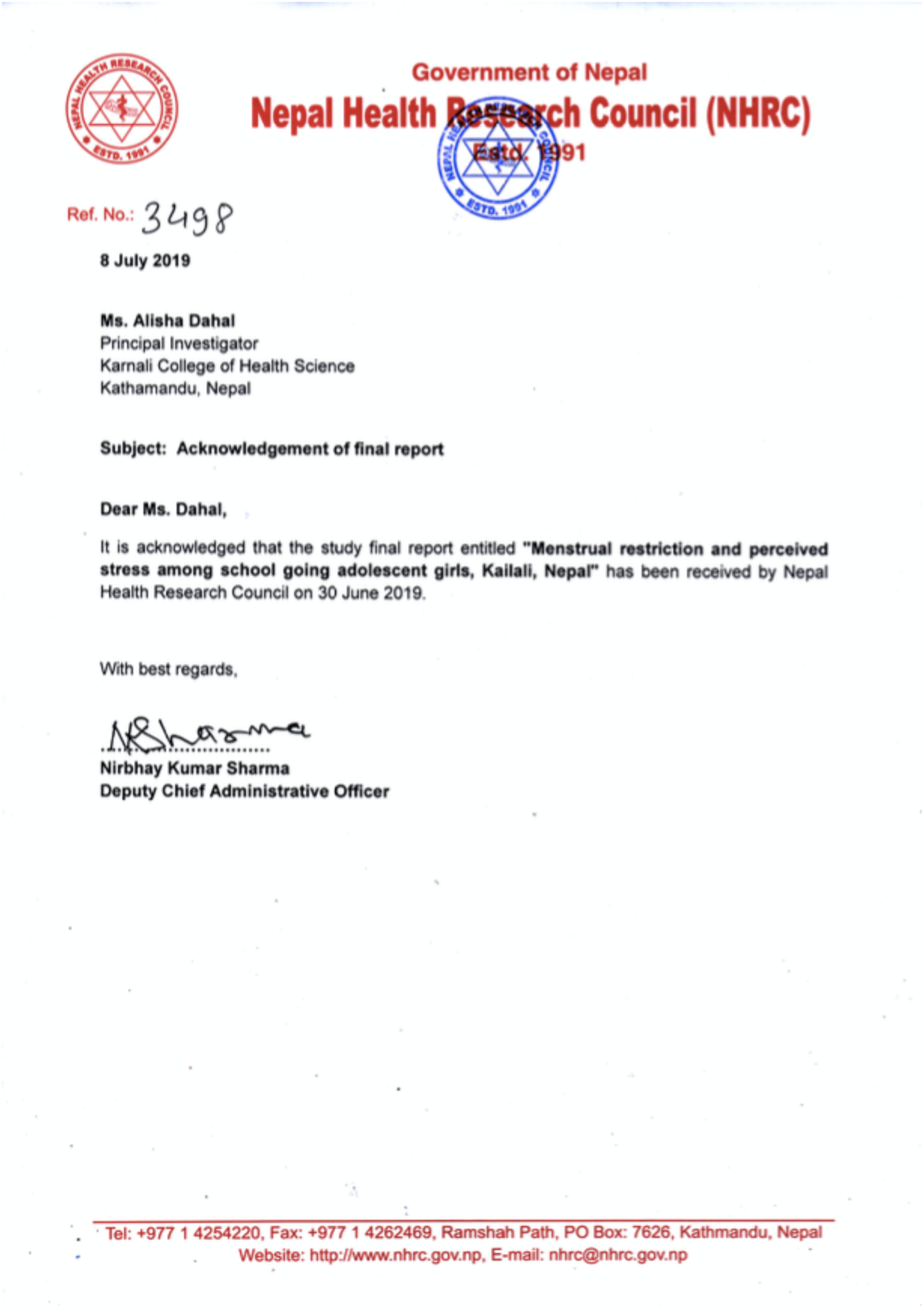

### Annex 6 Work Plan

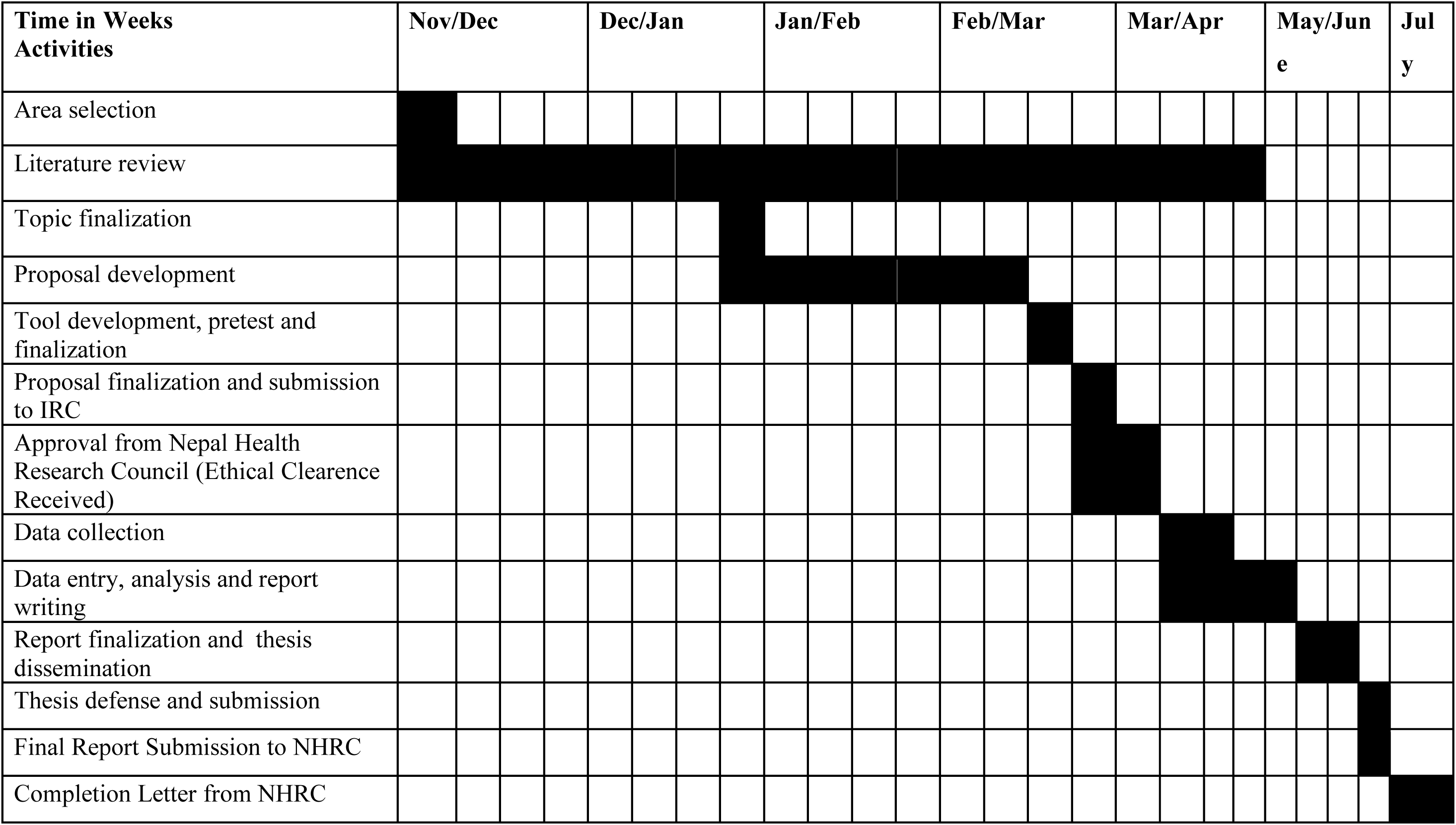

